# Costimulatory blockade depletes T peripheral helper, late-activated naïve, and DN2 B cells in rheumatoid arthritis

**DOI:** 10.64898/2026.03.14.26348386

**Authors:** Jasmine J. Shwetar, Abhimanyu Amarnani, William Rigby, Sladjana Skopelia-Gardner, Kelly V. Ruggles, Gregg J. Silverman

## Abstract

Rheumatoid arthritis (RA) is a chronic inflammatory autoimmune disease that causes joint destruction along with extra-articular morbidity and early mortality. Abatacept (CTLA-4 Ig), a blocker of lymphocyte co-stimulation, has become a well-accepted biologic treatment with proven efficacy in established-RA and for preventing disease onset in predisposed individuals. To investigate the immunologic implications of abatacept treatment, we conducted a prospective, open-label trial with multi-omic single-cell analyses of lymphocytes and BCR repertoire profiling at predefined intervals. Treatment-induced low-disease activity correlated with coordinated depletion of circulating peripheral helper cells (Tph), late-activated naïve cells (late-aNAV), and of CD27^-^IgD^-^ (Double negative, DN) Zeb2+CD11c+ T-box transcription factor 21 (Tbet^+^) DN2 unconventional memory B cells, implicated in the tertiary lymphoid structures responsible for the propagation of pathologic autoimmune responses and joint destruction. Among B-cell subsets, DN2 had the greatest representation of molecular machinery for antigen-uptake, processing, and presentation. Among memory B-cell subsets, DN2 had the lowest representation of somatically generated N-glycosylation sites and somatic hypermutation. Yet abatacept induced DN2 cells to express elevated CXCR4 levels, which normalized upon drug withdrawal, suggesting that abatacept treatment may cause these cells to traffic out of pathologic synovial infiltrates. In conclusion, we have documented that abatacept affects the circulating immune cellular drivers of disease activity, Tph, late-aNAV and DN2. Therapeutic depletion of these pathologic lymphocyte subsets is associated with clinical benefits that can persist after therapy cessation. Hence, levels of these subsets may serve as surrogates for the overall burden of disease and potential response to abatacept therapy.

**Graphical Abstract:** 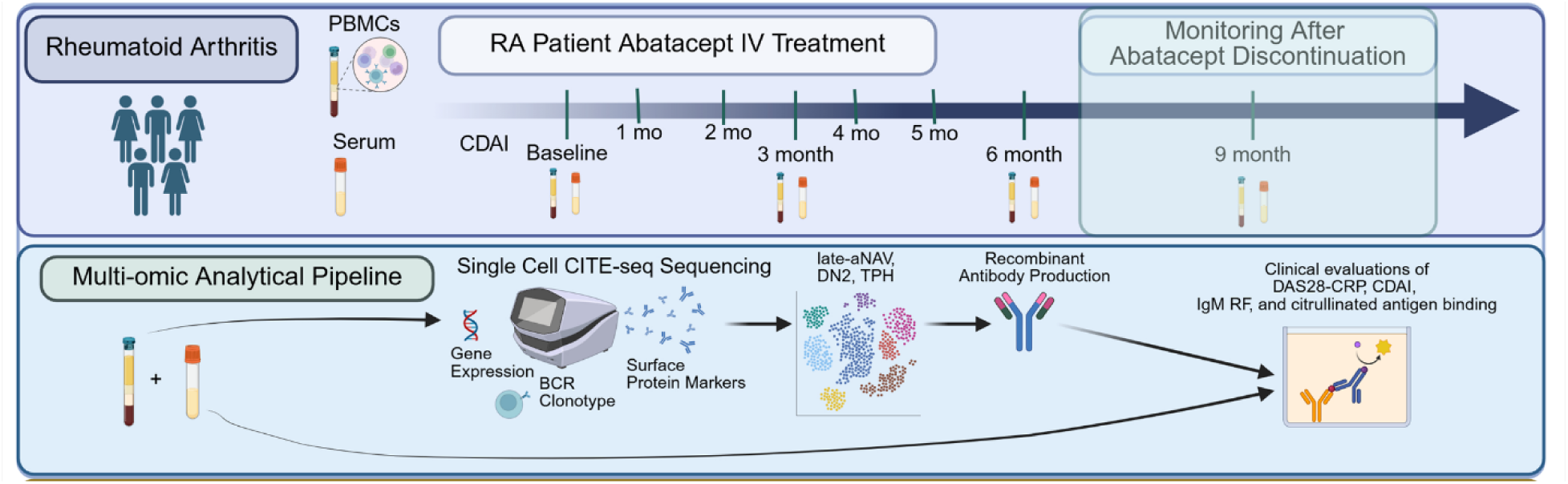

**One Sentence Summary:** Multi-omics analyses showed costimulatory blockade depletes trafficking DN2 B cells and Tph cells that correlates with rheumatoid disease response.

## INTRODUCTION

Rheumatoid arthritis (RA) is an autoimmune disease characterized by polyarthritis and progressive joint destruction, associated with severe extra-articular morbidity and early mortality (*1, 2*). RA affects approximately 0.5-1% of the population worldwide (*1, 3*), and its pathogenesis is complex (*4*). Disease initiation is postulated to begin with the activation of CD4+ T cells, a proinflammatory cascade of cell proliferation (*5*), and the induction of rheumatoid factors (RFs) and anti-citrullinated protein autoantibodies (ACPA), which in turn further contribute to downstream synovial inflammation (*4*).

The pathologic rheumatoid synovial lymphocytic infiltrates often form tertiary lymphoid structures (TLS), which drive articular and periarticular tissue destruction (*6, 7*). These TLS are, in part, populated by pathologic lymphocyte subsets with transcriptomic profiles that reflect dysregulated pathways of immune tolerance (*8, 9*). Perpetuation of this chronic condition depends entirely on the continuous trafficking of immune effector cells through the circulation, into and out of affected joints, and entry into previously unaffected ones (*10*).

Abatacept consists of the extracellular domain of human CTLA-4 fused to a modified Fc fragment of IgG, which is designed to interfere with the full lymphocyte activation that results from the binding of CD28 on T cells to CD80/CD86 on diverse types of antigen-presenting cells (APC)(*11–13*). Abatacept is an effective therapy in both patients with early RA and those with refractory disease who have had inadequate prior therapeutic responses (*4, 14*). In the AMPLE trial, abatacept demonstrated efficacy and safety equivalent to the TNF inhibitor, adalimumab (*15*), which disrupts downstream inflammatory pathways but does not affect the underlying pathogenic defect in immune tolerance (*16*). In contrast, abatacept interferes with a step hypothesized to be early in the immune pathogenesis of RA (*17*), and this agent reverses subclinical inflammation and attenuates disease progression and joint injury (*18, 19*). In controlled studies, in individuals at high risk of progressing from an asymptomatic state with circulating autoantibodies to active RA, abatacept reduced subclinical joint inflammation and significantly delayed progression to overt clinical disease (*20*) and improved inflammatory markers by up to 3 years (*21*). However, the broader effects of abatacept on pathogenesis remain poorly understood.

We therefore conducted a prospective, open-label, phase IV intervention trial in adult seropositive RA patients with moderate-to-high disease activity. At the six-month endpoint, 16 of 18 patients achieved low disease activity. To comprehensively survey the immune landscape (*22, 23*) in these RA patients, we performed quantitative single-cell analysis of blood samples obtained at predefined sequential time points, which were analyzed side by side to best characterize the time-dependent changes induced by abatacept (CTLA4-Ig). We integrated longitudinal single-cell transcriptomics and surface marker immunophenotyping with clinical trajectories in abatacept-treated RA patients. This multi-omic approach enabled us to characterize and quantify circulating T helper (Tph), late-activated naive (late-aNAV), and DN2 memory lymphocyte subsets that index abatacept response and effects on disease burden. Our analyses documented that abatacept treatment significantly reduced the levels of these trafficking pathologic lymphocyte subsets and normalized somatic alterations in the BCR repertoire, which correlated with high-hurdle, low-disease clinical benefits.

## RESULTS

### Clinical outcomes of Rheumatoid arthritis and Memory B Cells Trial (RAMBA)

In this open-label prospective study, four of the 25 initially identified patients were excluded as screen failures. The remaining 21 adult patients met the 2010 American College of Rheumatology/European League Against Rheumatism (ACR/EULAR) classification criteria for RA (*24*), and all had at least 3 tender and at least 3 swollen joints, as assessed by the 28-joint Disease Activity Score (DAS28) (*25*), at both screening and Day 1. All patients were serologically positive for both IgG anti-citrullinated protein antibodies (ACPA) and IgM rheumatoid factor (RF) by clinical laboratory testing. Inclusion required moderate-to-high disease activity, as defined by a Clinical Disease Activity Index (CDAI)(*26*) >16 and a DAS28-CRP >4.0 (*27, 28*).

The full course of abatacept infusions was completed by 18 patients (characteristics summarized in **Table 1**). At the predefined primary endpoint at month 6, 16 patients (89%) achieved low disease activity and were classified as responders (R). Following this assessment, if this low disease activity persisted, abatacept was discontinued for 4 additional months (through month 9 of the study). Flare after month 5, which was defined as DAS28-CRP >4.0 and/or CDAI >16, was documented in six patients (see **Table 1**) with a mean time to flare of 8 weeks after the last infusion (range: 3 to 11 weeks) (**Fig. 1**). The remaining 12 of 18 patients (67%) maintained remission without therapy until the end of the trial at month 9 (**Fig. 1A-D**). During the study period, no serious adverse events requiring hospitalization or intravenous antibiotic treatment occurred.

**Figure 1.**
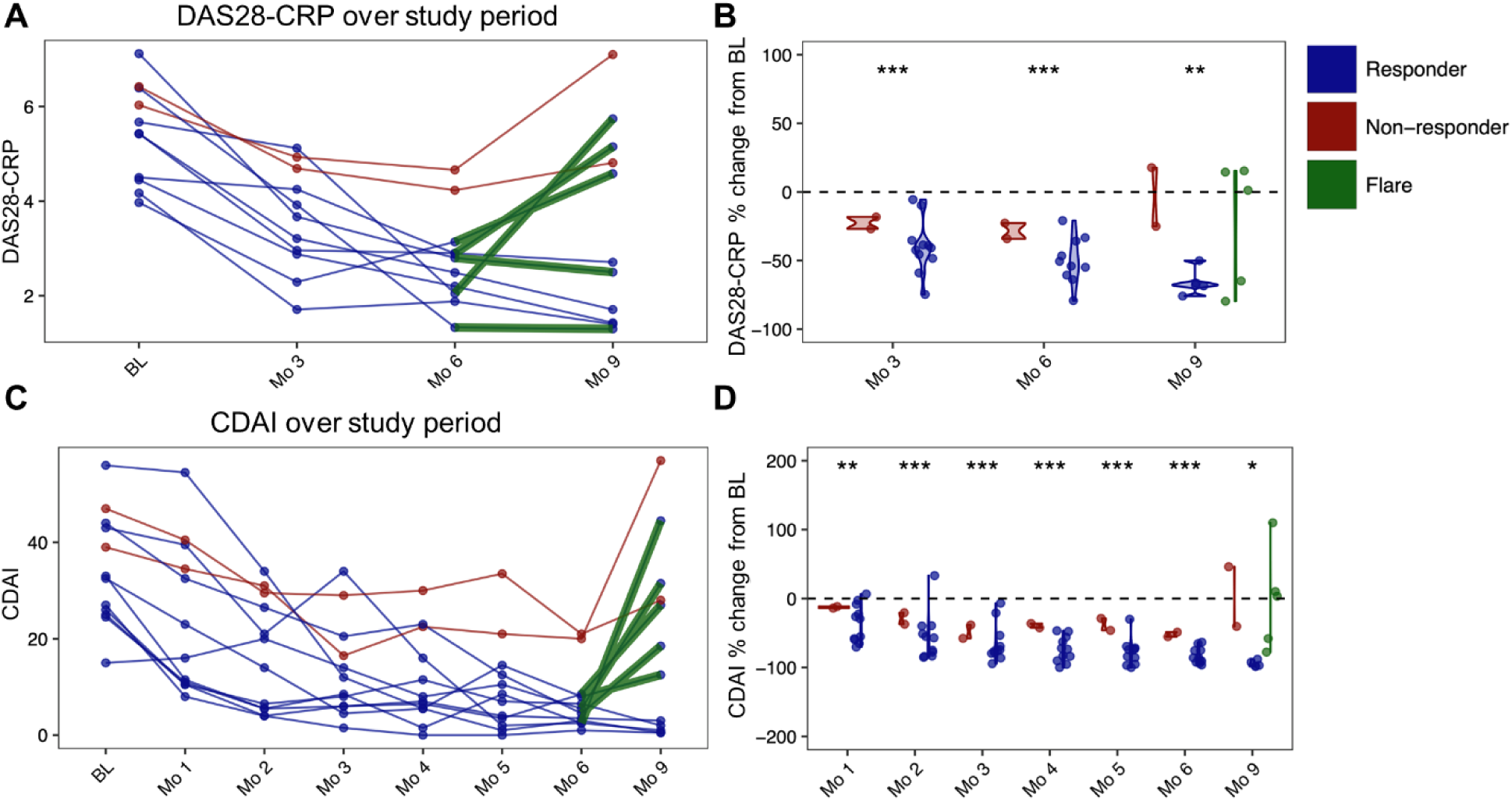
RA clinical disease activity scores pre, during, and post treatment with Abatacept. (**A**) Disease trajectories showing changes in DAS28-CRP scores over time for patients with complete data. Each line represents an individual patient’s disease course from screening through nine months of follow-up. (**B**) Percent change in Disease Activity Score 28 (DAS28) -CRP from screen. (**C**) Disease trajectories showing changes in Clinical Disease Activity Index (CDAI) scores over time for patients with complete data. Each line represents an individual patient’s disease course from screening through nine months of follow-up. (**D**) Percent change in CDAI from Screen. Significant changes from baseline are indicated by asterisks (* p<0.05, ** p<0.01, *** p<0.001). Dots represent individual patient data points.

**Table 1.** Patient characteristics.

| Characteristic | Value |
| --- | --- |
| Sex (M/F) | 14/7 |
| Age | 56.7 (25-72) |
| Methotrexate | 11/21 (52%) |
| Prednisone | 6/21 (29%) |
| Smoking Hx | 15/21 (71%) |
| Biologic naïve | 12/21 (57%) |
| RA duration >3 yrs | 12/21 (57%) |
| CDAI (SEM) | 34.5 (2.5) |
| DAS28-CRP (SEM) | 5.27 (0.19) |
| RAPID (SEM) | 13.8 (0.97) |
| Screen fail | 4/25 |
| Flare before 6 months | 3/21 |
| Completed 6 months | 18/21 |
| No flare at 9 months | 12/21 |
| Flare between 6 and 9 months | 6/21 |

Multi-omics analysis without fractionation for pts #13, 14, 17, 18, 21, 22, 23.
Multi-omics analysis with B cell enrichment for pts #1, 4, 9, 12, 19, 20.
Early treatment termination for pts 2,7, 8.
Patients with flares after 6 months (pts #1, 4, 11, 13, 21, 23) with a mean of 8 weeks after last infusion.
Patients who completed 9 months without a flare (pts #3, 5, 9, 10, 12,14, 16, 17,1819, 20, 22).

Among the 16 patients for whom complete blood samples were available, comparative subgroup analysis did not demonstrate statistically significant interactions between baseline factors, including disease duration, with changes in autoantibody levels and the primary outcome at 6 months. In addition, changes in IgG anti-citrullinated peptide (CCP3) autoantibody did not correlate with changes in CDAI or DAS28 clinical activity **(Table S1)**.

### T cell landscape and transcriptional reprogramming during abatacept treatment

To evaluate cellular and molecular changes induced by abatacept, we performed Cellular Indexing of Transcriptomes and Epitopes by Sequencing (CITE-seq) (*29*) on PBMC samples in patients with RA, which enables simultaneous quantification of single-cell transcriptomes and surface protein expression within the same cells. In these studies, we characterized the immune cell-surface proteins, single-cell transcriptional profiles, and B-cell receptor (BCR) repertoires (*29*) of peripheral blood mononuclear cells from 11 of the abatacept-treatment responders, and the two non-responders (NR) (patients #13 and #23) **(Table 1)**. In each experiment, we sequenced single cells from PBMC samples of an individual patient at four predefined time points: baseline (pre-treatment), 3 months of treatment, 6 months of treatment, and 3 months after cessation of therapy (month 9). At 3 months after treatment cessation, six of the responders **(see Table 1)** experienced disease flares at 9 months. Across the four time points, 55,497 high-quality T cells were captured from seven subjects.

Clustering identified eight canonical T cell subsets, including CD4 and CD8 naïve, T effector memory (TEM), Regulatory T (Treg), Mucosal Associated Invariant T (MAIT), Natural Killer (NK), and Proliferating T cell subsets (**Fig.2A, B**). To better understand the diversity within CD4 T cell responses to targeted inhibition of CD4 T cell co-stimulation by abatacept, and to identify functionally distinct CD4 subsets with potentially distinct treatment responses, we performed high-resolution clustering of CD4 TEM (**Fig.2C**). This identified four additional CD4 subsets: T helper 2 (Th2) defined by *GATA3* and *IL4R* expression, T helper 17 (Th17) defined by *RORA* and *CCR6*, Cytotoxic T helper 1 (Th1) with high expression of *RUNX3*, *STAT4*, and cytotoxic molecules (*GZMA*/*GZMK*), and lastly T peripheral helper (Tph), a pathologic T cell distinguished by its expression of *MAF*, *PDCD1*, and *HLA-DR* molecules (**Fig.2C, D**), which has been reported to be elevated in the circulation of patients with RA (*8, 30, 31*), and these circulating Tph are reported to be closely related to, and arise from stem-like Tph that reside in pathologic synovial infiltrates (*8*). In circulating PBMC samples from these patients, a distinct cluster of T follicular helper cells (Tfh) was not identifiable based on high CXCR5 levels, which are reported to maintain and define this T cell subset within lymphoid follicles (**Fig.S1**).

**Figure 2.**
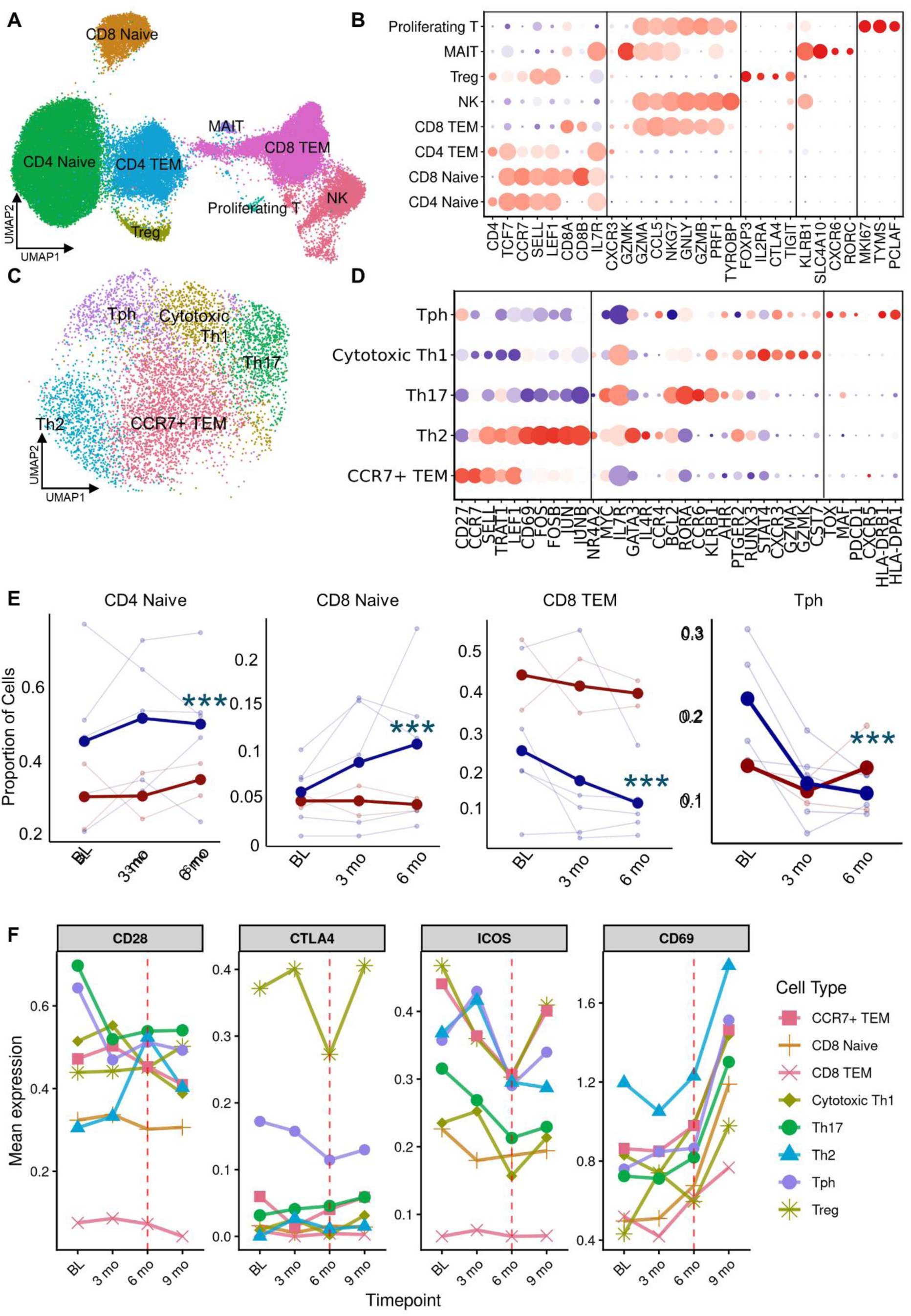
T-cell landscape in RA pre-, during, and post-treatment with Abatacept. (**A**) UMAP of T-cells from samples without B-cell enrichment, including Naïve, T Effector Memory (TEM), Regulatory T-cells (Treg), Natural Killer (NK), Mucosal Associated Invariant T-cells (MAIT), and Proliferating T-cells. (**B**) Defining markers for subsets in A. Dot size represents the percent expression. Dot size represents percent expression; Red denotes high expression, and purple denotes low. (**C**) UMAP of sub-clustering of CCR7+ TEM subsets, including T helper 2 (Th2), T helper 17 (Th17), Cytotoxic T helper 1 (Th1), and T peripheral helper cells. (**D**) Defining markers for subsets in C. (**E**) Changes in CD4 Naïve, CD8 Naïve, CD8 TEM, and Tph over time with responders in blue and nonresponders in red. Faded lines represent individual patients, and bold lines show the mean proportion for responders and nonresponders. The 9-month time point is not included due to lower cell recovery. Asterisks denote chi-square p-values comparing cell type proportions at baseline to 6 months in responders with p < 0.05 = *, p < 0.01 = **, and p<0.001 = ***. (**F**) Mean expression of co-stimulation genes CD28, CTLA4, ICOS, and CD69 in T cell subsets over time.

To understand changes in T-cell subsets associated with abatacept treatment, we compared representation of T cell subsets at baseline and at the 6-month primary endpoint. In responders, treatment increased both CD4 and CD8 naïve subsets over time, while CD8 TEM levels were reduced (**Fig.2E**). Other T cell subsets, including Treg, NK, MAIT, Th1, Th2, and Th17 populations, showed no significant changes between responders and non-responders (**Fig.S2A, B**). In all responders, total CD4 TEM abundance generally remained stable, whereas the Tph subset showed consistent and significant time-dependent decreases (**Fig. 2E**).

Analysis of genes encoding for T cell co-stimulatory molecules revealed treatment-dependent transcriptional changes. By month 6, CD28, which represents the primary co-stimulatory receptor targeted by abatacept (*11–13*), showed significant downregulation across all T cell subsets except Th2 cells. The most pronounced decreases were observed in Th17 and Tph cells (**Fig. 2F**). CTLA4, which encodes the inhibitory co-stimulatory receptor that competes with CD28 for CD80/CD86 binding, demonstrated subset-specific downregulation, most notably in Tregs followed by Tph cells at the primary endpoint. During treatment, expression of ICOS, an alternative co-stimulatory receptor, decreased across all subsets, with the greatest reductions in CD4+ T cells compared with CD8+ T cells.

Following cessation of abatacept, at month 6 the co-stimulatory molecule expression patterns began to revert, with a state of increased expression by month 9. Whereas both CTLA-4 and ICOS expression increased following drug withdrawal, CD28 expression showed more limited increases. In contrast, *CD69*, which represents an early cellular activation marker, exhibited a distinct temporal expression pattern. In fact, CD69 showed progressive upregulation across all T cell subsets, with the most pronounced increases occurring after treatment cessation between months 6 and 9 (**Fig. 2F**).

### Identification and characterization of functionally distinct B cell populations in RA

To deepen the detection of these CD19+ lymphocyte subsets, we enriched for B cells in samples from 13 patients at all time points; a total of 32,403 high-quality B-lineage cells were captured. Clustering identified pre-switch subsets that included transitional B cells, and three distinct naïve B cell subsets: resting naïve (rNAV), early activated naïve (early-aNAV), and late activated naïve (late-aNAV) (**Fig.3A**), which were identified based on both transcriptomic profiles and surface phenotype (**Figs.3B, C and S3**). In addition to antibody-secreting cells (ASCs), six distinct conventional memory B cell (MBC) subsets were also identified, characterized by high expression of the surface markers CD27 and CD21, along with differential expression of activation markers, including CD80, CD86, and CD95 (**Figs.3B** and **S3**). A small subset that clustered with post-switch B cells (which have undergone class-switch recombination to IgG, IgA, or IgE) was notable for lacking these surface markers but for high surface expression of CD11c and the CD19 B-cell lineage marker. This latter subset had a specific transcriptional signature, including *ITGAX, TBX21 and ZEB2* expression, which identified these cells as the double-negative 2 (DN2) subset of B cells (*32, 33*) (**Fig. 3C**), a disease-associated atypical memory subset with expansions reported to be T-cell dependent in patients with seropositive RA (*9, 33*) (as reviewed in (*34*)). Consistent with an earlier report (*33*), we observed that the transcriptomic profiles of the late-aNAV and DN2 B cells displayed considerable overlap (**Fig.3C**), with elevated surface protein identification of CD11c, a critical distinction in DN2, but not late-aNAV B cells (Fig. 3B,C) (*32, 33*). Whereas BCL2 family members regulate cellular survival through the expression of pro-apoptotic and anti-apoptotic proteins, DN2 B cells exhibited a characteristic balanced profile, with elevated anti-apoptotic members, *MCL1* and *BCL2A1* (also called Bfl-1 and *A1*), and co-expression of the pro-apoptotic genes *BAX* and *BID* (**Fig. S4**).

**Figure 3.**
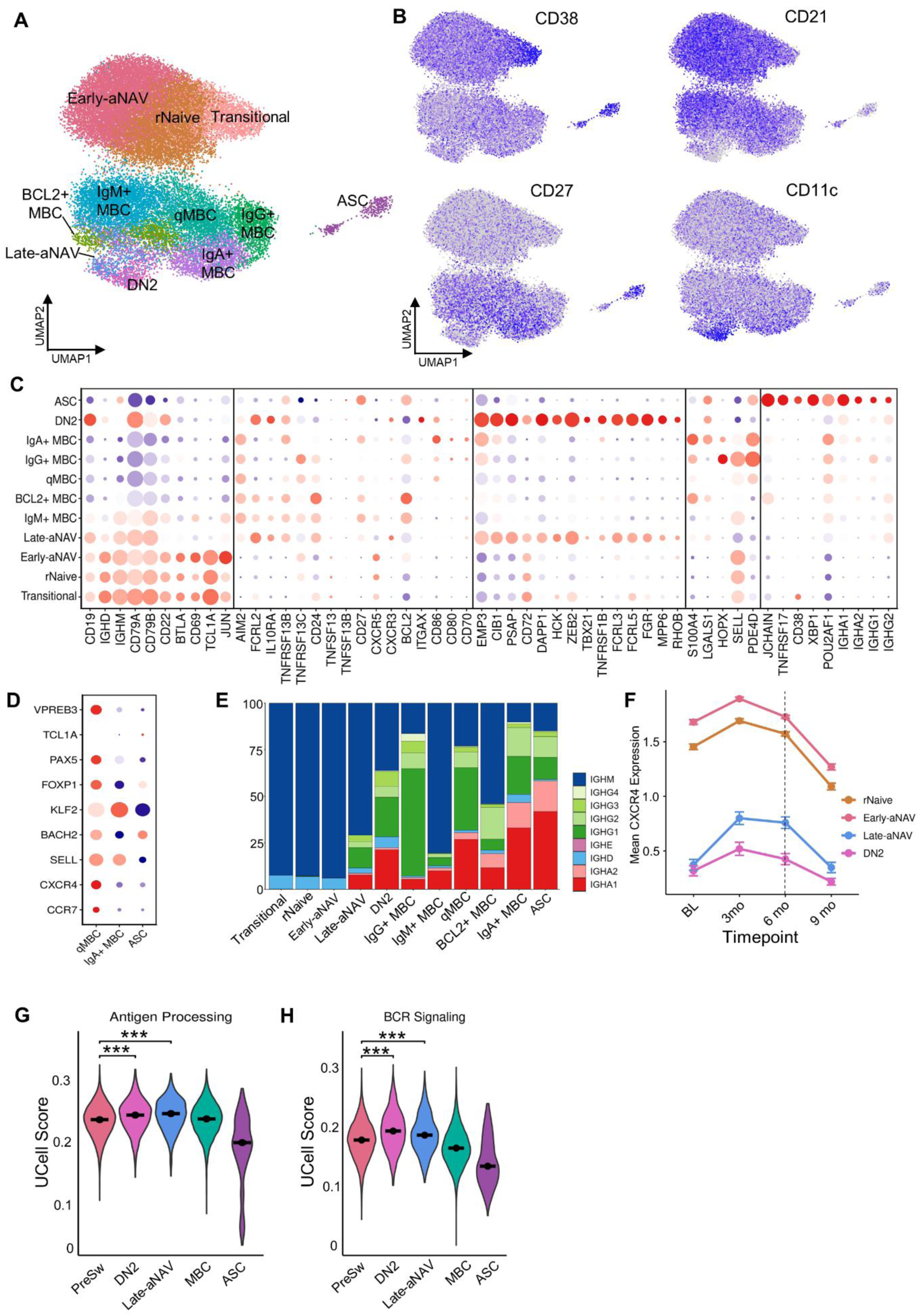
(**A**) UMAP of B-cells from samples including Transitional, resting Naive (rNaive), early activated naive (early-aNAV), late-aNAV, Double Negative 2 (DN2), Memory B-cells (MBC), and Antibody Secreting Cells (ASC). (**B**) Normalized expression of selected ADTs. (**C**) Defining transcription expression markers for subsets in A. (**D**) Defining transcription expression markers for qMBCs, IgA+ MBCs, and ASCs. Dot size represents percent expression; Red denotes high expression, and purple denotes low. (**E**) Cell types by heavy chain isotype and (**F**) CXCR4 expression in late-aNAV and DN2 B cells over time. **(G-H)** Module score of Antigen Processing & Presentation (KEGG pathways) and BCR signaling (KEGG pathways) pathways in PreSw, DN2, MBCs, and ASCs. PreSw B cells include all B cells that are transitional, rNaive, and early-aNAV.

At the transcript level, conventional MBC subsets had moderate CD27 and AIM2 expression (Fig. 3C). The IgG+ and IgA+ MBC subsets (**Fig. 3C**) had the highest expression of co-stimulatory molecules, CD80, CD86, and CD70, indicating their enhanced activation state (**Fig.3C**). Among the post-switch MBCs, *XBP1* and *ARID3A* emerged as standard regulators in both IgA+ and IgG+ MBCs (**Fig.3C**). As expected, ASCs showed strong dependence on XBP1 and POU2AF1, known regulators of plasma cell differentiation and immunoglobulin secretion (**Fig.3C**). Furthermore, two additional memory subsets exhibited distinct transcriptional profiles: a BCL2+ MBC subset characterized by high expression of pro-survival BCL2 as well as CD24 that is also linked to enhanced survival capacity (*35*). A quiescent MBC subset (qMBC) was marked by the expression of the transcriptional repressor, BACH2, and the pre-B cell receptor VPREB3, involved in p53-dependent apoptosis, as specific profiles that differ between the subsets within the memory B cell compartment (**Fig.3D**). In addition to RNA transcripts, MBC subsets were distinguished in part by the isotype association of their VDJ rearrangement transcripts (**Fig.3E**), highlighting a potential additional level of heterogeneity of their antibody products.

### DN2 B cells exhibit immune activation signatures and dynamic regulation of CXCR4

To assess time-dependent differential autosomal gene expression, we compared baseline to 6 months using linear mixed-effects models with random effects for responders **(Table S2).** Across B cell subsets, we identified 12 genes with significant time-by-cell type interactions **(Table S2).** Among these, *CXCR4 (*p = 0.021) ranked 6th of 138 genes. Significant treatment-induced *CXCR4* expression increases were found from baseline to month 6, across all four B cell subsets, which was most prominent in late-aNAV cells. Furthermore, from month 6 to month 9 after abatacept cessation there were substantial decreases in CXCR4 (**Fig.3F**). This pattern of temporal changes was very similar in resting naïve, early-aNAV, as well as the related late aNAV and DN2 subsets (**Fig.3F**). These findings suggest there may be treatment-dependent modulation of this chemokine receptor, which regulates B cell trafficking and lymphoid node retention. By blocking trafficking into pathologic synovial sites, changes in CXCR4 expression could therefore contribute to abatacept-induced clinical benefits by potentially redirecting these lymphocytes into physiologic lymphoid organs, where these pathologic effector functions may be dissipated.

Given the marked treatment-responsive decrease in DN2 representation and the associated late-aNAV, we used pathway-scoring analysis to further characterize the potential intracellular pathways expressed in these subsets (**Fig. 3G-H**). DN2 cells had significantly higher scores for antigen presentation and processing (APC activity) and for BCR intracellular signaling pathways than all other B cell subsets, including pre-switch populations, conventional MBCs, and ASCs. These activities may be intertwined in membrane associated BCR cross-linking interactions via autoantigens, which drive autoimmune pathogenesis by internalizing, processing, and presenting peptides from autoantigens.

To understand the developmental relationships between B cell subsets, we performed trajectory inference analysis using transitional B cells, which are emigrants from the central compartment, as the root population. Pseudotime ordering revealed differentiation paths that begin with transitional cells, then progress through naïve B cell state toward memory and effector populations, and include distinct paths toward MBCs, or late-aNAV and DN2 B cells (**Fig. S5A, B**). Spatial autocorrelation analysis using Moran’s I statistic (*36*) identified genes with coordinated changes in expression for long trajectories. These included transcriptional signatures at the distinct differentiation branches of early-aNAV, late-aNAV/DN2, and conventional memory B cells (**Fig. S5C-F**).

### Treatment-induced changes in defined circulating lymphocyte subsets

For the B cell subsets that precede these stages of maturation, there was a significant time-dependent increase in the combined pre-switch population (transitional, rNaive, and early-aNAV subsets) by the primary endpoint in responders **(Fig. 4A)**, with the temporal dynamics of each individual subset shown in **Fig. S6A-C.** This accumulation of earlier developmental stages could be explained as a consequence of abatacept-induced arrest of their differentiation into the late-aNAV and DN2 subsets. Responders also showed numerical decreases in the representation of CD27+ MBCs, although these changes did not attain statistical significance (**Fig. 4B**; individual memory subset dynamics in **Fig. S6D-H**). Most notably, responders demonstrated a highly significant decrease in the combined DN2 and late-aNAV population by the primary endpoint (p < 0.001) **(Fig. 4C)**, whereas ASC proportions remained stable in responders **(Fig. 4D)**.

**Figure 4.**
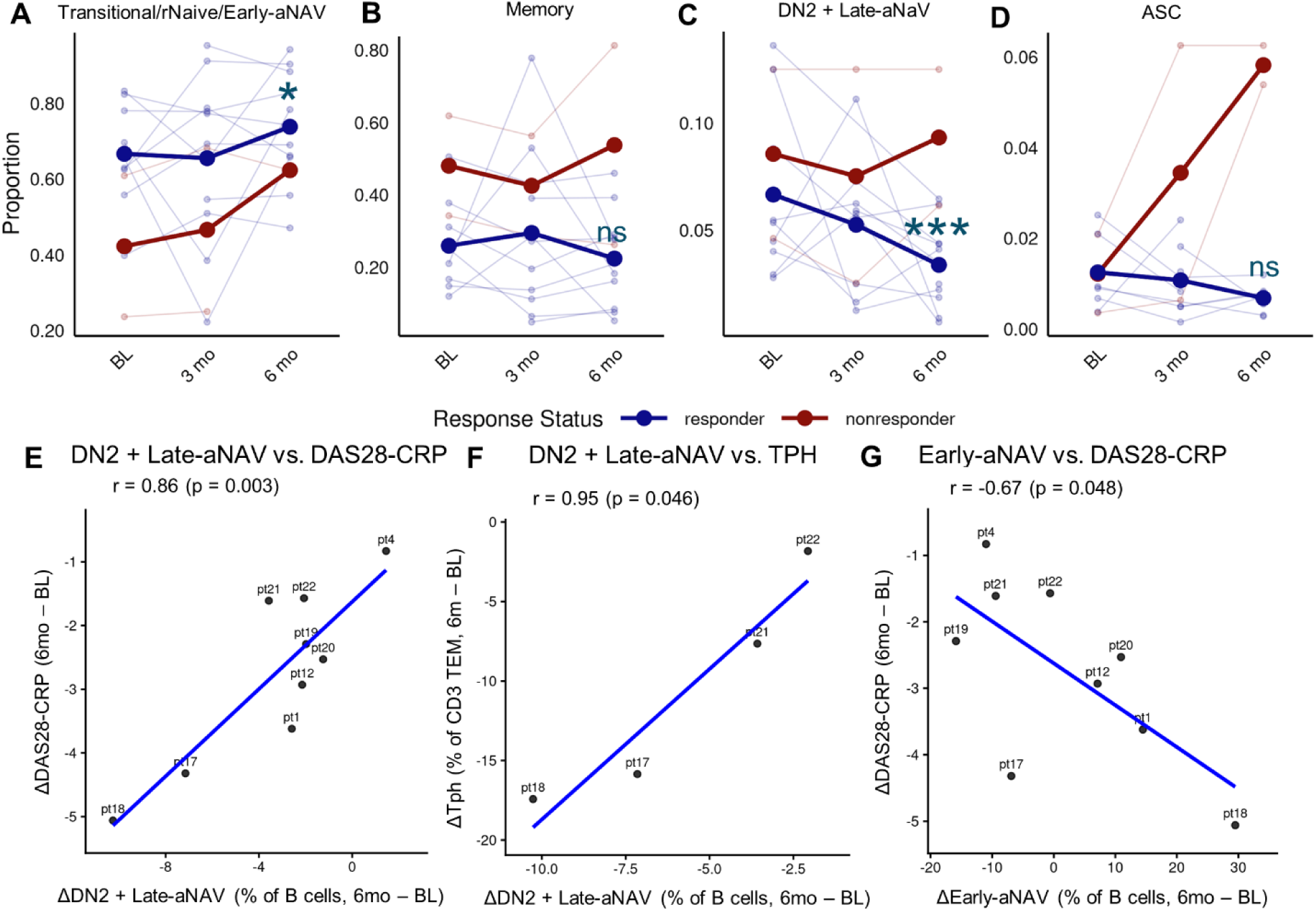
Changes in B cell states and clinical outcomes with Abatacept treatment and cessation. (**A-D**) Mean proportion of Pre-switch (Transitional, rNaive, and early-aNAV), Memory (all MBC subsets), DN2 + late-aNAV B cells, and ASCs across time for responders and non-responders. Faded lines represent individual patients, and bold lines show the mean proportion for responders and nonresponders. Asterisks denote chi-square p-values comparing change in proportion of B cells at 6 months vs. baseline in responders with p<0.05 = *, and p<0.001 = ***. Linear regression of change in proportion of DN2 and late-aNAV over treatment period vs change in (**E)** DN2 + Late-aNAV vs. Disease Activity Score (DAS28-CRP), **(F)** DN2 + Late-aNAV vs. Tph, and **(G)** Early-aNAV vs. DAS28-CRP.

### Treatment-induced changes in disease-associated subsets correlate with clinical disease remission

To further investigate the clinical relevance of the pronounced treatment-dependent decreases in abatacept responders, we examined correlations between the representation of B cell subsets and clinical disease activity. Improvement in disease outcomes (DAS28-CRP(*25*) and CDAI(*26*)) had a strong direct correlation with a decrease in late-aNAV together with DN2 B cells for DAS28-CRP (r = 0.86, p = 0.003, **Fig. 4E**), and for CDAI there was a similar, but not significant, numerical trend (r = 0.61, p = 0.084, **Fig.S7**), with a consistent pattern across all responders. Extended analysis across all B cell subsets confirmed that these associations were specific to late-aNAV and DN2, but not other subpopulations (**Fig. S7**). When late-aNAV and DN2 B cell subsets were grouped together, over time there was a direct coordinated concurrent decrease in abundance of Tph (r = 0.95, p = 0.046) (**Fig. 4F**). In contrast, the early-aNAV subset instead was negatively correlated with DAS28-CRP (r = -0.67, p = 0.048) (**Fig. 4G**). Indeed, this correlation was more pronounced than in any other lymphocyte subset (**Fig. S7**). Our findings, therefore, indicate a specific association between the B cells implicated in extrafollicular responses and the Tph subset of T cells, which are the reciprocal arms of the adaptive immune system and are intertwined in these pathologic responses (*37*).

Our trial included only two non-responders, limiting our ability to perform meaningful statistical comparisons. However, as with responders, non-responders also showed time-dependent increases in pre-switch B cell populations, including transitional, rNaive, and early-aNAV. Yet, we observed distinct trends in B cell dynamics for other subsets (**Figs. 4A, 4C**). In contrast to responders, however, non-responders showed trends toward modest increases in conventional MBC subsets, particularly IgG+ and IgA+ memory populations, although these trends did not reach statistical significance (**Fig. S6**).

Most strikingly, while responders demonstrated significant treatment-induced decreases in DN2 B cells (p < 0.001), this trend was reversed in the non-responders, who instead showed modest increases in Late-aNAV and DN2 frequency at the primary 6-month endpoint (**Fig. 4C**). Furthermore, in responders, ASC proportions remained stable throughout treatment, whereas non-responders exhibited marked expansion of this effector population, with ASC frequencies more than doubling from baseline to 6 months (**Fig. 4D**).

### Abatacept alters the signature of somatic mutations in the B cell repertoire of memory subsets

To assess the influence of therapy on the immune repertoires expressed in these B cell subpopulations, we examined patterns of somatic hypermutation (SHM), and somatically generated N-glycosylation sites, both indicators of the influence of T helper cells and clonal selection. As expected, at baseline, transitional and naïve B cells had little or no evidence of SHM. Notably, among all memory populations, DN2 cells exhibited the lowest SHM frequency, suggesting limited experience with T cell help (**Fig.S8A, B**). DN2 cells also displayed a low level of somatically generated N-glycosylation sites, in contrast to the higher levels in the conventional memory subsets and ASCs (**Fig.S8A, B**). Longitudinal analysis revealed treatment-dependent changes in the prevalence of N-glycosylation motifs in the late-aNAV and ASC populations (**Fig.S8C, D**). Among these conventional memory populations, IgA+ MBCs demonstrated the highest SHM burden (**Fig.S8A**), and ASCs exhibited the greatest frequency of N-glycosylation sites (**Fig.S8B**). In contrast, both SHM and somatically-acquired N-glycosylation sites were few or absent from newly generated transitional and resting naïve B cells (**Fig.S8A, B**).

### An expanded DN2 B-cell clone with a characteristic autoreactivity pattern

BCR repertoire analysis of individual cells was used to identify B-cell clones with two or more members (**Fig. S9A**). In Patient 1, we identified a markedly expanded clone (clone 1) (**Fig. S9A**), comprising 19 cells that appeared to derive from the same antibody rearrangement events, and the daughter cells were distributed across late-aNAV B cells (n=2), DN2 B cells (n=4), and BCL2+ memory B cells (n=13) (**Fig. S9B**). All cells in this clone expressed IgM BCR rearrangements. Notably, this clone was identified in a patient who was a responder to abatacept at 6 months, and after treatment cessation, this patient experienced a disease flare at month 9. Longitudinal tracking demonstrated that this clone persisted throughout the treatment course, with detection at all 4 sampling time points.

Sequence analysis of clone 1 identified a VH rearrangement of the VH3-30*04, DH6-13*01, and JH3*02 germline segments, with somatic hypermutation responsible for seven silent and six replacement mutations, with only one located in a complementarity-determining region (CDR). The light chain showed the closest homology to Vκ3-15*01 and J1*01, with one silent and five replacement mutations, of which one was in a CDR subdomain within the encoding antibody gene (**Fig. S10,11**).

To functionally characterize this clone, a recombinant IgM-antibody was generated **(Fig. S12)**. This antibody exhibited prominent binding reactivity for the Epstein-Barr Virus (EBV) EBNA-1 protein (**Fig. S13A)** and was also reactive with the synthetic cyclic citrullinated peptide (CCP3) used in RA diagnosis(*24*). This antibody showed substantially reduced binding to the non-citrullinated control peptide (CQP3), confirming its fine specificity for citrulline (**Fig. S13B,C**). The polyreactivity of this autoantibody was shown through binding of nucleosomes, native DNA and insulin. Antibodies against citrullinated EBNA-1 peptides may cross-react with citrullinated fibrinogen in patients with ACPA-positive RA(*38*). This is clinically relevant, as RA patients generally display elevated levels of EBNA-1 IgM antibodies, as do patients with other autoimmune diseases(*39*). These antibodies are prevalent in patients with circulating rheumatoid factors and ACPAs (*40, 41*).

A germline-reverted variant lacking somatic point mutations compared to the closest known germline genes showed substantially reduced binding to EBNA-1, CCP3 and other autoantigens, indicating that somatic hypermutation enhanced autoreactivity (**Fig. S12, S13**). This antibody may therefore be an example of clonal selection of a B cell for anti-viral responses, that was also recruited into poly- autoreactive responses during RA pathogenesis. This supports the theory that EBV contributes to the generation and diversification of pathogenic autoantibody responses that target (citrullinated) self-antigens in patients with seropositive RA. This pattern therefore also aligns with recent evidence demonstrating that EBV infection may reprogram autoreactive B cells, a paradigm now established across multiple systemic autoimmune diseases (*39*).

Collectively, these data demonstrate that clonal expansion of somatically mutated, polyreactive B cells within the DN2 compartment can give rise to autoantibodies targeting citrullinate-modified antigens implicated in RA pathogenesis. The persistence of this clone throughout abatacept treatment, and the representation of daughter cells across multiple B-cell subsets suggest that such clones may represent an important therapeutic target in RA.

## DISCUSSION

In this prospective study, longitudinal single-cell peripheral blood profiling yielded several key findings regarding the effects of abatacept on patients with RA. Firstly, in treatment responders, we observed significant treatment-induced compositional changes in both T- and B-cell compartments, with time-dependent increases in naïve T cells and pre-switch transitional, resting, and early-aNAV B cells. Abatacept also induced contractions in levels of circulating Tph, and in the late-aNAV and DN2 B-cell subsets, which paralleled clinical remission. In most patients, these treatment-induced subset changes persisted after drug withdrawal without disease activity rebound.

Secondly, we found striking correlations between key cellular and clinical parameters, with reductions in late-aNAV and DN2 strongly correlating with clinical improvement as measured by DAS28 (r = 0.86) and with changes in Tph cell frequency (r = 0.95). Indeed, consistent with treatment-induced clinical benefits, we found that naïve T cells increase, while Tph levels decrease. These findings align with studies showing that abatacept modulates T cell activation and differentiation in patients with autoimmune disease. In RA, Fukuyo et al. reported that T follicular helper-like (Tfh-like) cells were significantly reduced after abatacept therapy (*42*). While we sought also to track effects on Tfh, findings for circulating Tfh in RA patients have been conflicting (*43*). When we used CXCR5 in clustering studies, we did not identify a distinct circulating CD4+ T cell subset [**Fig. S1**].

Thirdly, at the molecular level, we documented that DN2 B cells have the highest expression of genes involved in antigen uptake, processing and presentation (**Fig . 3G-H**). This B cell subset has a distinctive gene signature, including *ITGAX (CD11c)*, *FCRL3*, *FCRL5*, *HCK*, and *FGR*, that is regulated by transcriptional networks involving *TBX21*, *IRF5*, *TCF4*, and *KLF12*. These findings position DN2 B cells not only as autoantibody precursors but also as professional antigen-presenting cells that perpetuate T cell-driven autoimmune responses within the rheumatoid synovium.

Finally, we characterized a representative persistent polyreactive B cell clone distributed across the DN2 and other memory compartments in a clinical responder who experienced a disease flare following withdrawal of abatacept treatment. This clone provided direct evidence that these cellular populations express BCR well-suited for autoantigen uptake, processing and presentation to disease-associated T cells, possibly including the Tph that may also play roles in disease recurrence. Our findings, therefore, link abatacept’s mechanism of action to patient clinical trajectories and nominate circulating Tph and DN2 signatures as candidate cellular biomarkers of disease burden and treatment durability that could inform decisions on therapeutic continuation vs. tapering, and/or discontinuation.

Among all B cell subsets, DN2 and the related late-aNAV subset exhibit the most pronounced abatacept treatment-dependent changes in blood representation, further supporting their central roles in disease. Unlike conventional memory B cells, DN2 cells have reduced thresholds for differentiation into plasma cells and are associated with extrafollicular responses (*9*). From our studies, we observed that the DN2 cell subset can be associated with broad autoreactivity and a low frequency of somatic hypermutation in expressed antibody rearrangements. These findings are consistent with a cellular origin outside of conventional germinal centers — which may represent extrafollicular pathways within ectopic lymphoid structures — where defects in clonal negative selection may permit escape from immune tolerance (*44*). These characteristics likely reflect DN2’s dependence on T-cell help for survival and function, which explains their sensitivity to co-stimulation blockade in clinical responders.

Our findings document that DN2 B cells and the related late-aNAV are critical downstream mediators of abatacept treatment response, and these findings contribute to evolving perspectives on the surface phenotype and potential functional roles of these cells (*45*). The molecular pathway scores of DN2 cells strongly support their roles in professional antigen presentation and processing, with transcriptional regulation by TBX21 driving T-bet-mediated inflammatory responses and by IRF5 promoting interferon-γ signaling pathways. These cells were initially characterized by their heightened responsiveness to TLR7 stimulation and readiness to differentiate into plasma blasts (*33*). Previously, DN2 B cells were found to be expanded in patients with systemic lupus erythematosus (SLE) with active disease and nephritis, with phenotypic, functional, and transcriptomic profiles similar to, and in many ways overlapping with those of late-aNAV B cells (*33*). Wing et al reported that DN2 B cells are the major precursors of synovial ASCs in patients with RA (*9*). Our studies suggest that the efficacy of abatacept in RA may partly arise from its ability to disrupt this pathogenic differentiation pathway.

The correlated reductions in Tph and DN2 populations suggest a functional relationship that may be central to RA pathogenesis and treatment response. The strong correlation between treatment-associated reduction of the DN2/late-aNAV subset and clinical improvement (DAS28-CRP, r = 0.86, p = 0.003) (**Fig. 4E**) directly links these cells to synovial disease activity. In contrast, there is an inverse correlation between the level of Early-aNAV and change in DAS28-CRP between baseline and 6 months of treatment (r= -0.67, p=0.048) (**Fig. 4G**), which indicates these cells increase with treatment. Strikingly, an even stronger correlation between DN2 and Tph cell dynamics (r = 0.95, p=0.046) (**Fig. 4F**) suggests abatacept may disrupt the cellular interaction required for their continued survival. This Tph-DN2 relationship is further supported by reports that the capacity of DN2 cells to differentiate into plasma cells and subsequently produce autoantibodies is driven by IL-21 (*33*), a key cytokine produced by Tfh and Tph cells (*30, 46*). Providing a mechanistic link between our observations in blood and the pathologic processes occurring within affected joints, Masuo et al. recently demonstrated that stem-like Tph residing in rheumatoid synovium give rise to effector Tph that circulate peripherally [8]. The dynamic regulation of CXCR4 in DN2 cells, which increased during treatment and then decreased after drug cessation, suggests a treatment-dependent modulation of this chemokine receptor, which regulates B-cell trafficking and retention within germinal center structures (*47*).

Notably, in contrast to responders, the non-responders showed increased representation of circulating ASC, without the depletion of DN2 cells, further indicating that B cell compartment dynamics may serve as early predictors of treatment response and durability. Analysis of the BCR repertoire identified a persistent, poly-autoreactive clone, assignable to the DN2 and MBC subsets, that recognizes the synthetic CCP3 peptide, as well as nucleosomes and native DNA. This finding aligns with other studies demonstrating that DN2 cells share clonal relationships with antibody-secreting cells (*48*), suggesting a dynamic developmental continuum. The persistence of this clone throughout treatment, despite overall DN2 reduction, highlights the challenges of eliminating pathogenic B cell responses and may provide a cellular correlate for post-treatment disease recurrence. The polyreactive nature of this clone is consistent with the hypothesis that DN2 B cells serve primarily as antigen-presenting cells rather than solely as precursors to highly specific autoantibody-secreting plasma cells, thereby linking autoantigen capture to T cell activation in a feed-forward loop.

Our findings demonstrate that abatacept fundamentally reprograms the immune landscape in patients with RA. There was a coordinated downregulation of transcripts for costimulation (*CD28*, *CTLA-4*, and ICOS) across T-cell subsets, which was most pronounced in the Th17 and Tph populations (**Fig 2F**), reflecting the drug’s direct effect on pathogenic T-cell subsets that drive RA inflammation (*31, 49*). Following drug cessation, co-stimulatory gene expression (*CD28*, *CTLA4*, ICOS) subsequently normalized (**Fig. 2F)**. Yet, during treatment there was a progressive increase in CD69 expression that accelerated dramatically after treatment cessation (**Fig. 2F**), which in some patients may contribute to disease recurrence. The increase in naive T- and B-cell populations in clinical responders further suggested that co-stimulation blockade by abatacept prevents these cells from receiving the CD28-mediated second signal required for full activation and differentiation into pathogenic effector subsets (*50*).

We documented that clinical improvement, as measured by DAS28-CRP disease activity metrics, was paralleled by progressive reductions in circulating disease-associated T-cell and B-cell subsets. However, while single-cell RNA analysis pinpoints the representation of pathologic lymphocyte populations, peripheral blood markers may not mirror findings in the synovium, as extensive literature in rheumatoid arthritis (RA) has described the heterogeneity of synovial tissue pathology and the limitations of traditional disease activity scores like DAS28 in capturing this complexity. Furthermore, only a minor subset of rheumatoid synovial biopsies show a lymphocyte-predominant infiltrate. Hence, tertiary lymphoid organs (TLO) have been detected in only a minority of rheumatoid synovial biopsies (*51–54*). In this small trial, at baseline, these seropositive RA patients generally had elevated levels of trafficking Tph and DN2 (**Fig. 4**). If these lymphocytes are commonly the dominant pathologic drivers of seropositive RA and arise in TLO, the true prevalence of functional TLO may have been underestimated, in part due to issues arising from inadequate or incomplete biopsy sampling.

Our findings may also be relevant to understanding how abatacept blocks progression from pre-clinical RA to clinically active RA. Tph and DN2 B cells have also been implicated in the earliest stages of autoimmune amplification, before clinical symptoms manifest (*5*). In the ARIAA and APIPPRA trials, abatacept delayed or prevented disease onset in at-risk individuals with circulating autoantibodies but no clinical arthritis (*20, 21*). Our data suggest that the mechanism underlying this preventive effect likely involves disruption of the Tph-DN2 cellular partnership before tertiary lymphoid structures become established in synovial tissue. By interrupting T-B collaboration at this early checkpoint, abatacept may prevent the self-perpetuating cycle of autoantigen presentation, autoantibody production, and inflammatory amplification that characterizes the transition to clinical disease and tissue injury.

Several limitations should be considered when interpreting our findings. This was a single-center, small, open-label study, which may limit the generalizability of our results. Also, our analysis was limited to peripheral blood samples, as no synovial biopsies were performed; hence, concurrent immunological changes within the synovial infiltrates were not captured.

However, our observations are consistent with randomized controlled trials examining autoantibody responses to abatacept. Additionally, all patients in our study were double autoantibody-positive (ACPA+ RF+), which may represent a distinct subgroup, which is particularly responsive to abatacept treatment, potentially limiting extrapolation to seronegative RA patients. Lastly, while our transcriptomic analysis provides valuable insights into cellular responses to treatment, functional studies will be needed to further confirm the mechanistic relationships between synovial infiltrating lymphocytes and the changes in trafficking lymphocytes that were the focus of the current studies.

Using longitudinal blood-based single-cell data, we have inferred an integrated model of abatacept’s effects on Tph and DN2 programs that track with disease activity in RA. During active disease, autoantigen presentation in the ectopic lymphoid tissues of affected joints creates a specialized microenvironment in which the stem-like Tph cells provide help to aNAV and DN2 B cells in the lymphoid synovial infiltrates (*8*), thereby driving autoantibody production and inflammatory responses. Co-stimulation blockade inhibits Tph cell activation, which we postulate severs the T-B cellular interactions required for DN2 B cell maintenance and survival. The remarkably tight correlation between Tph and the DN2/late-aNAV reductions (r = 0.95), and the strong association between reductions of DN2/late-aNAV cells with clinical benefits (r = 0.86) support this mechanism. DAS28, an accepted surrogate continuous metric for clinical disease activity, showed a strong direct correlation with the representation of lymphocyte subsets by single-cell analysis. These data therefore support our 2014 hypothesis regarding the profound effects of abatacept on RA-associated autoimmune memory (*55*), and deepen our understanding of the varied contributions of different B cell subsets to RA pathogenesis (*56*).

During treatment, we observed the progressive upregulation of CXCR4 specifically in DN2 B cells, and we postulate that these potentially pathogenic cells are then redirected out of inflammatory sites toward secondary lymphoid organs, where they may undergo further regulation or elimination. However, the persistent polyreactive B cell clone identified in a patient who experienced disease flare demonstrated that pathogenic B cells can nonetheless survive throughout treatment and subsequently drive disease recurrence once co-stimulation signaling is restored.

In conclusion, our study provides novel insights into the cellular and molecular mechanisms underlying abatacept’s therapeutic efficacy in RA, identifying trafficking DN2 B cells as key pathogenic mediators and their partnership with Tph cells as a central axis in disease perpetuation. The ability to predict treatment durability through molecular signatures present before treatment may also offer new opportunities for personalized therapeutic approaches.

These findings not only advance our understanding of RA pathogenesis but also suggest that targeting the DN2-Tph axis may be a promising strategy for achieving durable remission in patients with RA. Future studies should validate these biomarkers in larger cohorts and explore therapeutic interventions specifically designed to disrupt this pathogenic cellular partnership.

## MATERIALS AND METHODS

### Clinical and demographic characteristics and selection criteria

This study was approved by the NYU Langone and Dartmouth Hitchcock IRB. All patients signed informed consent. Patients (n=25) who fulfilled 2010 ACR-EULAR criteria for the RA diagnosis(*24*), and were double-positive for ACPA and RF by clinical testing, were considered eligible for enrollment for this phase IV open-label clinical trial, Rheumatoid <u>a</u>rthritis <u>M</u>emory <u>B</u> Cells and <u>A</u>batacept (RAMBA) (NCT03652961), conducted at the Dartmouth-Hitchcock Medical Center. Eligible patients were men or women aged 18 years or older who were not pregnant or nursing. All had active RA satisfying the ACR/EULAR 2010 classification criteria (*24*), with symptoms present prior to screening. Written informed consent was obtained before enrollment.

For enrollment, each RA patient had at least 3 tender and 3 swollen joints on the DAS28 joint exam at screening and Day 1, and at least moderate disease activity (CDAI >16; DAS28-CRP ≥4.0), indicating an inadequate response to methotrexate. Four of the 25 patients screened did not meet the inclusion criteria for disease activity and were deemed screen failures. More than half of the patients (57%) were naive to biologic DMARDs, and recruitment included patients who had previously received a TNF inhibitor and had discontinued it at least 5 months before screening. All subjects were also naive to rituximab, tocilizumab, sarilumab, targeted synthetic DMARDs such as tofacitinib, baricitinib, and upadacitinib, as well as investigational therapies for RA. Subjects were permitted to receive oral corticosteroids at a stable dose of ≤10 mg prednisone daily for at least 4 weeks. Subjects could not receive an IM, IV, or IA administration of a corticosteroid within 4 weeks before the screening visit or initiation of therapy.

Inclusion required at least moderate disease activity as defined by Clinical Disease Activity Index (CDAI) >16 or DAS28-CRP >4.0 (*27*). Four of the 25 patients were excluded as screen failures; the remaining 21 patients (14 men, 7 women) met the 2010 American College of Rheumatology/European League Against Rheumatism (ACR-EULAR) classification criteria for RA (*24*). All patients had at least 3 tender and at least 3 swollen joints, as assessed by the Disease Activity Score in 28 joints (DAS28) at both screening and Day 1, and were serologically double-positive for anti-citrullinated protein antibodies (ACPA) and rheumatoid factor (RF) by clinical laboratory testing.

A total of 18 patients with moderate-to-severe disease activity (mean DAS28-CRP of 5.27 and mean CDAI of 34.5) completed the full course of abatacept infusions (patient characteristics summarized in Table 1). At enrollment, the mean age was 57 years, and all were either intolerant to or had an inadequate response to methotrexate; 52% remained on weekly methotrexate. Six subjects were on daily monotherapy with other medications (HCQ (#1, 8, 9, 10, 13); with one receiving sulfasalazine (#2); while only four subjects were not receiving a daily oral corticosteroid dose (#5, 12, 18, 23).

Patients were dosed intravenously based on body weight (500 mg for those <60 kg, 750 mg for those 60-100 kg, and 1000 mg for those >100 kg). Abatacept was administered on Day 1, week 2, and then monthly at months 2-5, followed by de-challenge and subsequent follow-up at months 6 and 9, or earlier if a flare supervened. Patients were phlebotomized at screening, on day 1 (when they began abatacept therapy), at subsequent infusions at week 2 and months 2 through 5, and at the time of study exit if due to a clinical flare. Phlebotomy was performed for routine clinical laboratory studies and for sample isolation and cryopreservation at -80 ^°^C or in liquid nitrogen. As described above, abatacept was discontinued after the 5th month infusion. For patients who had achieved low disease activity by DAS28-CRP (<3.2) or CDAI (<10), abatacept was then held for up to 4 additional months (month 9) or until a flare occurred. A flare was defined by an examination demonstrating a DAS28-CRP greater than 4.0 or a CDAI >16.

### Multiplex serum autoantibody immunoassays

To evaluate the effect of abatacept treatment on disease-associated autoimmune responses, a validated multiple-bead-based (MBI) assay platform was used (*57*). Herein, this system was used to detect IgM- to human IgG Fc-fragments, and IgG-responses to a range of citrullinated peptides, run side-by-side with non-citrullinated peptide versions. For each patient, we compared antibody levels in baseline (screen, pretreatment) and 6-month (the primary endpoint) samples, with additional samples included when available. Each assay was performed with serum diluted 200-fold, 800-fold, and 3200-fold in duplicate. These assays included the CCP3 peptide used in the commercial assay (Inova Diagnostics). A standard RA serum pool was used for internal calibration across studies, with the activity index expressed as relative units (RU).

### Single-cell library preparation and sequencing

All samples were initially cryopreserved under careful conditions using the protocol from the Immune Tolerance Network, and later thawed and then analyzed by cell indexing of transcriptome and epitopes by sequencing (CITE-seq) (*29*), using a modified protocol with sample multiplexing using hashtags (HTOs), surface phenotyping using antibody-derived tags (ADTs), and 5’ BCR sequencing. In the same experiment, all PBMC samples from different time points obtained from a single patient were run side by side, along with the sample from the same healthy adult control. In each of seven experiments, samples from individual patients (pts #13, 14, 17, 18, 21, 22, 23) (Table 1) were compared and processed without enrichment. In six other experiments, the samples from an individual subject (pts #1, 4, 9, 12, 19, 20) were first subjected to magnetic bead-mediated negative selection (B cell enrichment kit 2, Miltenyi). Individual experiments included at least 50,000 hashed cells, processed with an emulsifier and the Titanium platform (10X Genomics).

### Alignment, preprocessing, and quality control

Sequencing data was processed using the Cell Ranger Multi pipeline (v7.0, 10x Genomics) to generate gene expression matrices, ADT count matrices, HTO count matrices, and V(D)J repertoire data. We used the Seurat R package v5 for data analysis (*58*). For quality control, we filtered cells with fewer than 200 unique genes or more than two standard deviations above the mean gene count in a sample. Additionally, cells with more than 30% of reads mapping to mitochondrial genes were removed. Samples were demultiplexed using Seurat’s HTODemux function with centered log-ratio (CLR) normalization. This allowed us to assign cells to their corresponding sample origins and retain only singlet cells, as scDblFinder was used to remove remaining doublets.

Following normalization, variable feature identification, and principal component analysis with Seurat’s NormalizeData, FindVariableFeatures, and RunPCA, respectively, batch effects were corrected using Harmony integration across each sequencing run (*59*). Unsupervised clustering was performed using the Louvain algorithm on a shared nearest neighbor graph constructed from the harmony-corrected principal components. Non-B-cell clusters were removed based on canonical marker expression at the transcript and surface protein level. Cell clusters were characterized using differential expression analysis. For sub-clustering of B and T cells, the same approach was applied. Notably, the 9-month time point was excluded from compositional analyses of T cell subsets because of low cell counts following successful hashtag demultiplexing, which would otherwise bias relative abundance estimates for rare cell populations.

### Statistical and computational analyses

B cell receptor sequences generated from the Cell Ranger V(D)J pipeline were processed using the Immcantation software suite (*60*). Raw V(D)J sequences were annotated using AssignGenes.py with IgBLAST (*61*) and converted to AIRR-compliant format with MakeDb.py. Sequences were filtered to include only productive, full-length, and high-confidence contigs.

Using the Scoper R package (*62*), clonal groups were defined based on identical V and J gene usage, identical CDR3 length, and nucleotide similarity in the CDR3 region, with a distance threshold determined by the distToNearest and findThreshold functions from the shazam package (*63*). Germline sequences were reconstructed using createGermlines, and somatic hypermutation (SHM) was quantified based on observed mutations. BCR isotype distribution was analyzed across cell types, time points, and patient response groups using the alakazam package (*64*). Clonal expansion and B cell lineage analyses were performed with the Dowser R package to track clonal evolution over time (*65*). Using maximum-likelihood methods, phylogenetic trees for selected clones were constructed in Dowser and assessed for evidence of measurable evolution across time points. To evaluate changes in B cell subset proportions between baseline and the 6-month time point in responder patients, Chi-square tests were used. Differential expression analysis between treatment time points was performed using Seurat’s FindMarkers function, applying the Wilcoxon rank-sum test with a Bonferroni correction for multiple comparisons.

### Recombinant monoclonal antibody production and antigen binding assays

To functionally characterize expanded B-cell clones of interest, recombinant monoclonal antibodies were generated from paired heavy and light chain BCR sequences identified through the single-cell V(D)J analysis. For Clone 1, the somatically hypermutated heavy chain (IGHV3-30*04/IGHD6-13*01/IGHJ3*02) and kappa light chain (IGKV3-15*01/IGKJ1*01) variable region sequences were codon-optimized and cloned into IgM constant region expression vectors (NYU 5-12). A corresponding germline-reverted variant (NYU8) was generated by reverting all somatic hypermutations in both the heavy and light chain variable regions to their predicted germline sequences, as reconstructed by the createGermlines function in the Immcantation suite using the IMGT reference directory. Recombinant antibodies were expressed by transient transfection of HEK293 cells and purified by Protein A affinity chromatography (Sinobiological). Purity was confirmed by SDS-PAGE. Antigen binding specificity was assessed using a multiplex bead-based immunoassay (MagPix/Luminex). Antibodies were tested for reactivity against a panel of antigens including cyclic citrullinated peptide 3 (CCP3), the corresponding native arginine-containing control peptide (CQP3), and nucleosomes. Results are reported as Net Median Fluorescence Intensity (Net MFI). Non-ACPA clonotype-derived recombinant antibodies from the same patient were included as negative binding controls.

Multiple sequence alignments of clonal daughter BCR sequences against germline gene segments were generated using SnapGene, with immunoglobulin region boundaries (FWR1–FWR4, CDR1–CDR3) annotated according to the IMGT numbering system(*66*), where CDR3 is defined as positions 105-117 in the IMGT numbering system, spanning from the residue after the conserved Cysteine (C104) at the 3’ end of the V-segment to the residue before the conserved Tryptophan (W118, heavy chain) or Phenylalanine (F118, kappa light chain) at the 5’ end of the J-segment.

## Data Availability

All data are available in the main text or the supplementary materials, and databases are available upon reasonable request.

## Supplementary methods

There were no serious events requiring admission or intravenous antibiotics. Ten URI events were reported by eight patients, two of whom were confirmed as Covid-19 infections. One of these infections was reported at the month 2 visit, and symptoms had fully resolved by the month 3 visit. In a different patient, a COVID-19 infection occurred 9 months after resuming monthly abatacept; 3 months later, the patient fully recovered and resumed abatacept. Neither of these COVID-19 infections required hospital admission or anti-viral therapy. One patient was treated with azithromycin. In addition, there was one episode of dyspnea prompting an emergency room evaluation for underlying COPD. Two episodes of short-lived, nonspecific rash were also reported, with etiology never established. No episodes of Herpes Zoster were reported.

## List of Supplementary Materials

Fig S1. T follicular helper (Tfh) cells do not form a distinct cluster in peripheral blood CD4+ T effector memory cells from patients with rheumatoid arthritis.

Fig. S2. Mean proportion of T cell subsets.

Fig. S3. Normalized expression of antibody-derived tags across B cell subsets.

Fig S4. BCL family member gene expression across B cell subsets.

Fig. S5. UMAP visualization, trajectory analysis, and Moran’s I spatial autocorrelation analysis of B cell differentiation.

Fig. S6. Mean proportion of B cell subsets over time.

Fig. S7. Changes in late-aNAV and DN2 B cell abundance correlate with clinical outcomes and peripheral helper T cell dynamics.

Fig. S8. Somatic hypermutation levels and N-glycosylation motif prevalence across B cell subsets.

Fig S9. Phenotypic and clonal characteristics of expanded Clone 1

Fig S10. Nucleotide and amino acid sequence alignment of Clone 1 B cell receptor (BCR) heavy chain sequences.

Fig S11. Nucleotide and amino acid sequence alignment of Clone 1 B cell receptor (BCR) light chain (Igκ) sequences.

Fig S12. Amino acid sequence alignment of recombinant antibodies to germline

Fig S13. Antigen binding specificity of recombinant monoclonal antibody NYU5-12 (Clone 1) and germline-reverted antibody NYU8.

Supplemental Table 1.

Supplemental Table 2.

## Acknowledgements

We thank Sergei Koralov, Deepak Rao, Shiv Pillai and Scott Jenks for key advice.

We appreciate the contributions of our clinical coordinators and patients, and Zakia Azad and Jaren Buliyat for their technical assistance. We thank our colleagues Sheila Kelly and Michael Maldonado for their input during the work.

## Author contributions

Conceptualization: GJS, WFR

Methodology: GJS, WFR, AA, JJS, KR

Investigation: GJS, WFR, JJS, SS-G, AA

Visualization: JJS, GJS, AA

Funding acquisition: GJS

Project administration: GJS, WFR

Supervision: GJS, WFR

Writing – original draft: JJS, KR, GJS

Writing – review & editing: JJS, AA, KR, GJS, WFR, SS-G

