## Supplemental Figures for "Costimulatory blockade depletes T peripheral helper, late-activated naïve, and DN2 B cells in rheumatoid arthritis"

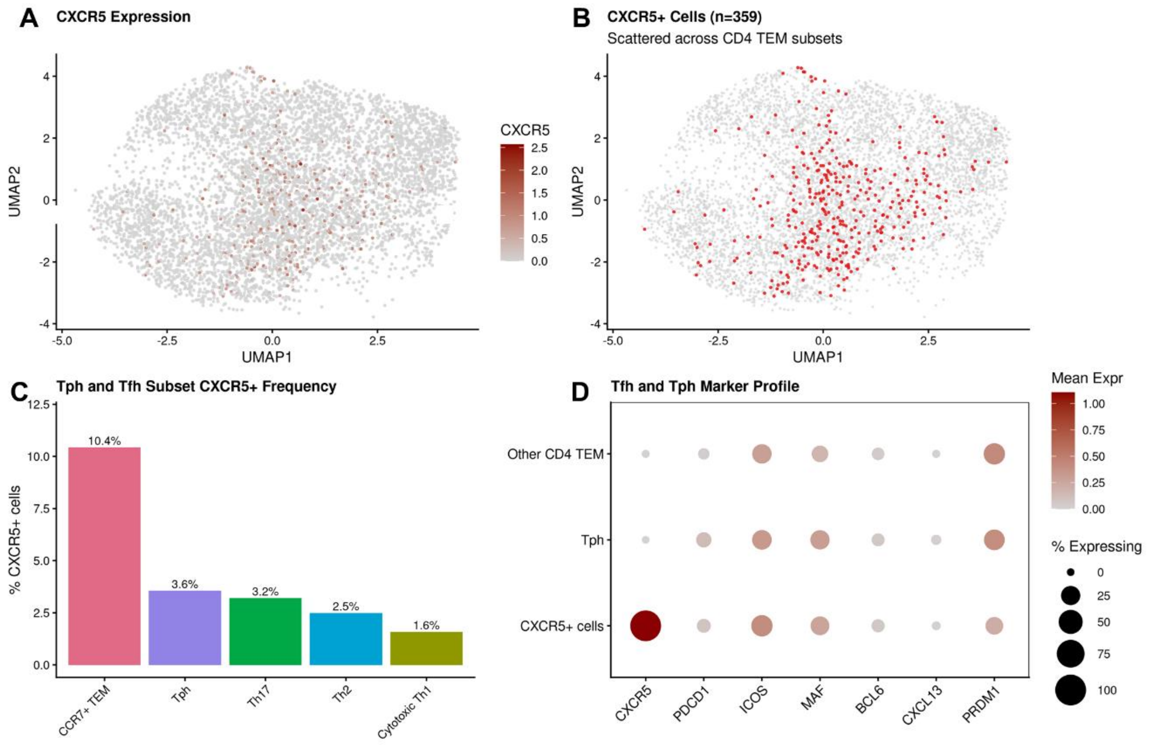


**Supplemental Figure 1.** T follicular helper (Tfh) cells do not form a distinct cluster in peripheral blood CD4+ T effector memory cells from patients with rheumatoid arthritis.
**(A)** UMAP visualization of CD4+ T effector memory (TEM) cell subsets identified in peripheral blood mononuclear cells (PBMCs) from rheumatoid arthritis (RA) patients. **(B)** UMAP projection showing CXCR5 expression levels across all CD4+ TEM cells, with color intensity representing normalized expression values. **(C)** Distribution of CXCR5+ cells (n=359) across the UMAP space, demonstrating scattered localization rather than a distinct transcriptional cluster. **(D)** Percentage of CXCR5+ cells within each CD4+ TEM cluster, with IER CD4 TEM showing the highest proportion (14.4%). Therefore, CXCR5+ cells are rare and do not form a transcriptionally distinct population.

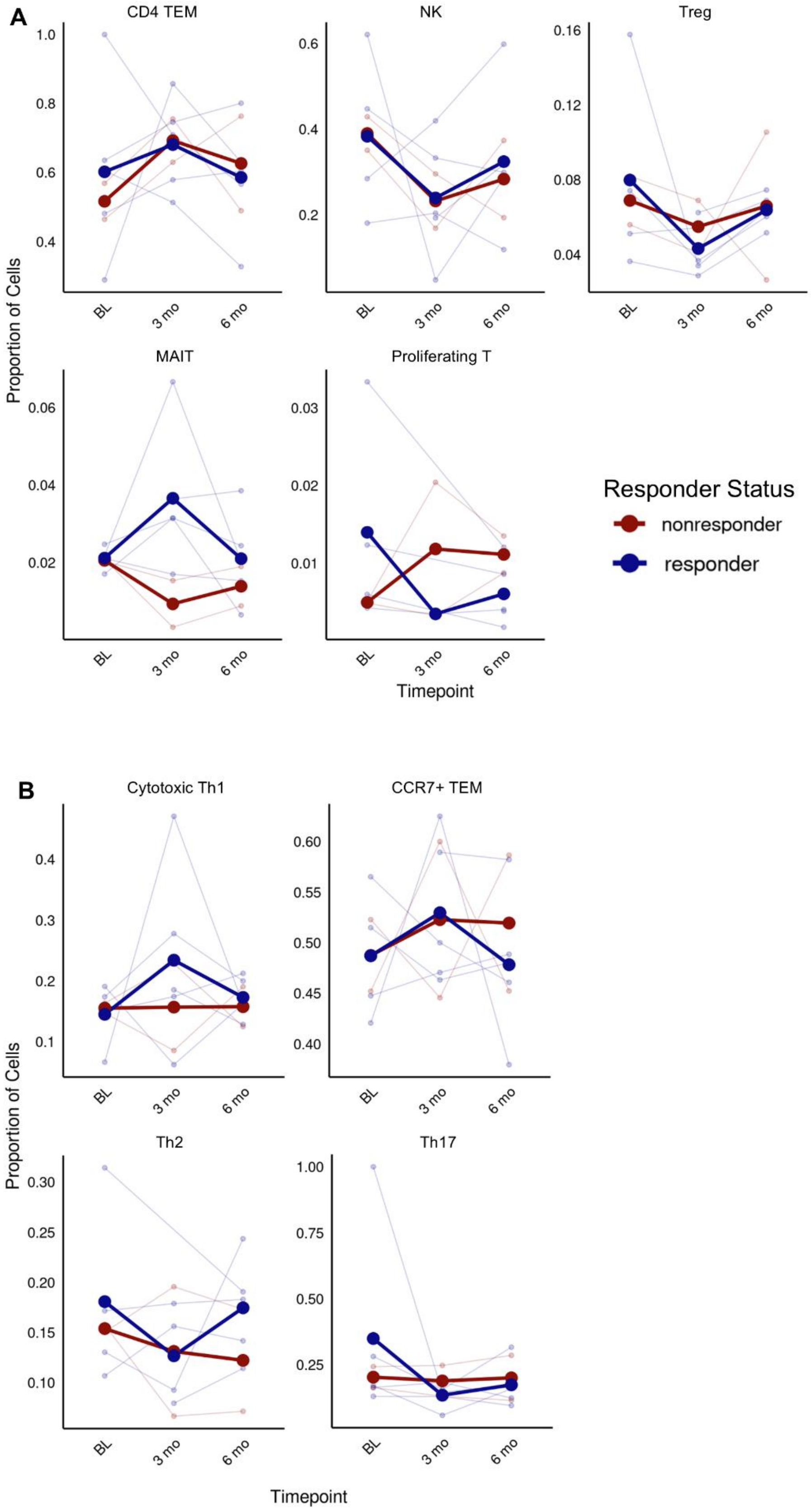
A
CD4 TEM
NK
Treg
MAIT
Proliferating T
Proportion of Cells
Timepoint
BL
3 mo
6 mo
Responder Status
nonresponder
responder
B
Cytotoxic Th1
CCR7+ TEM
Th2
Th17
Proportion of Cells
Timepoint

**Supplemental Figure 2.** Mean proportion of T cell subsets. (A) Mean proportion of T Effector Memory (TEM), Regulatory T-cells (Treg), Natural Killer (NK), Mucosal Associated Invariant T-cells (MAIT), and Proliferating T-cell subsets. **(B)** Mean proportion of Cytotoxic T helper 1 (Th1), CCR7+ TEM, T helper 2 (Th2), and T helper 17 (Th17) subsets over time. No significant differences were identified by chi-square comparing cell type proportions between responders and non-responders at 6 months.

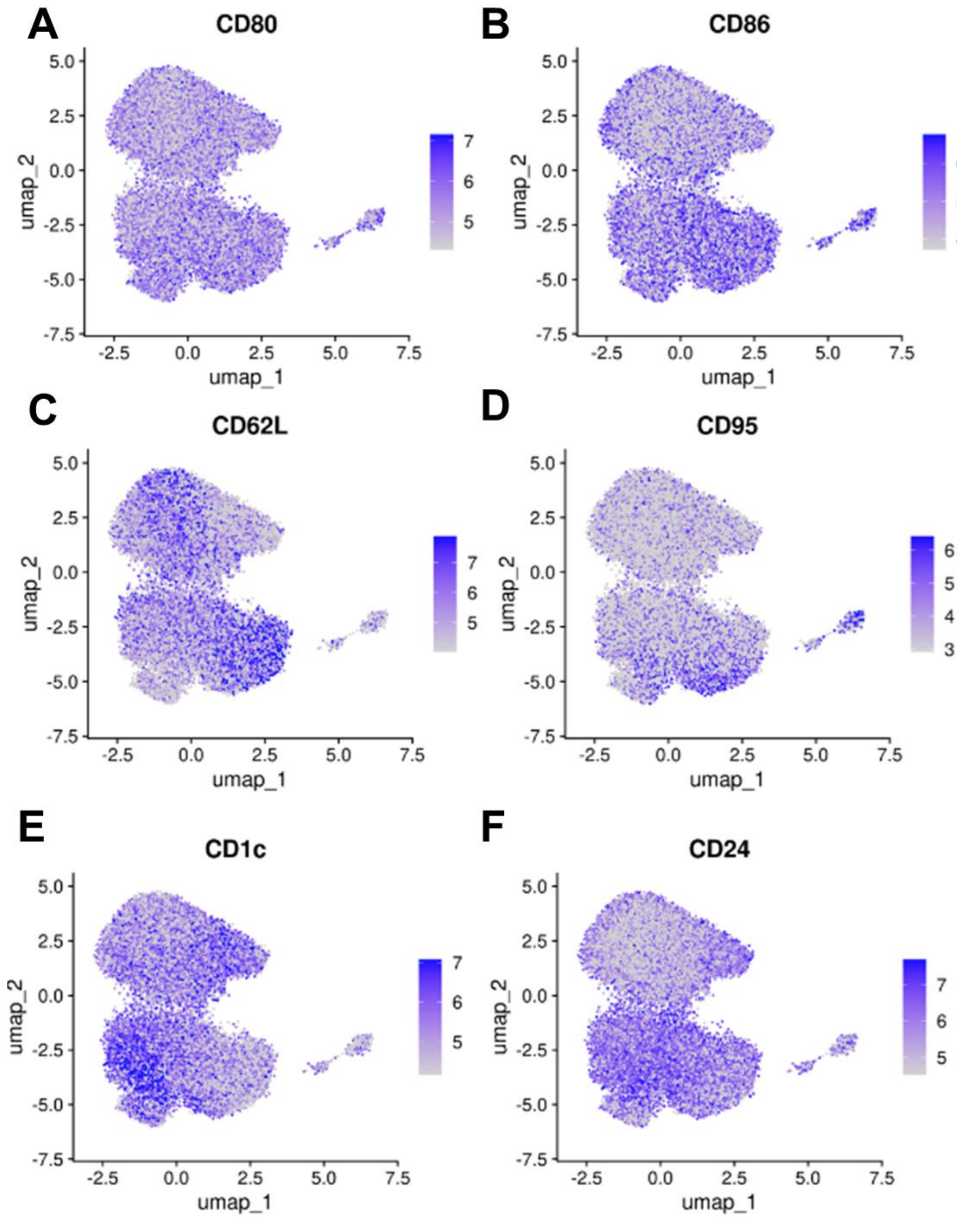


**Supplemental Figure 3.** Normalized expression of ADTs across B cell subsets related to Figure 3A-B. UMAP including **(A)** CD80, **(B)** CD86, **(C)** CD62L, **(D)** CD95, **(E)** CD1c, **(F)** CD24.

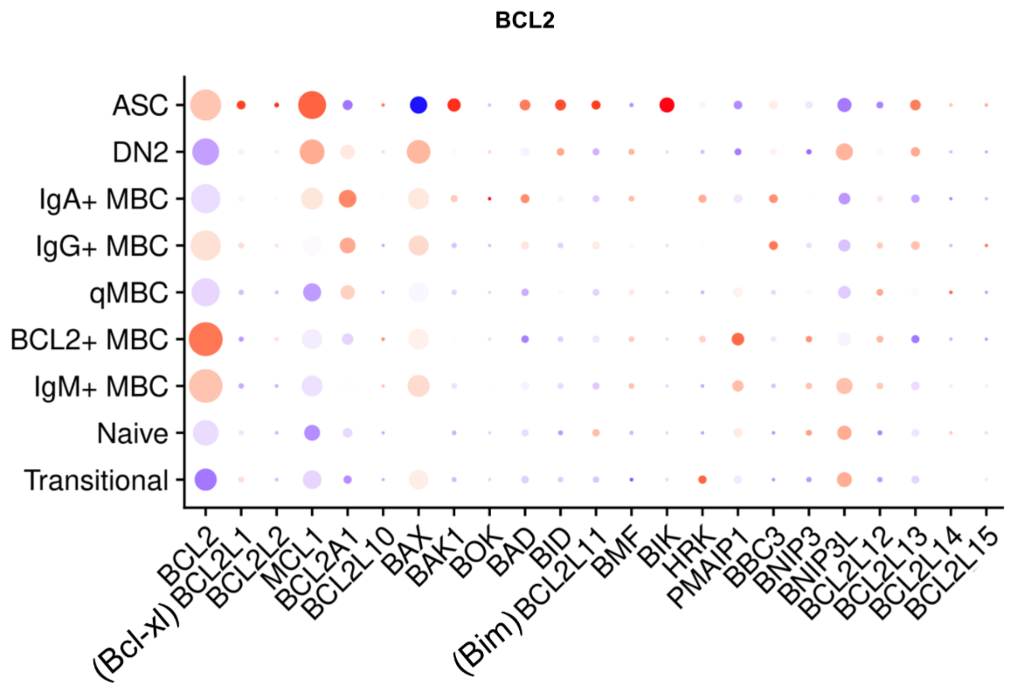


**Supplemental Figure 4.** BCL family member gene expression across B cell subsets. Dot size represents percent expression; Red denotes high expression and purple denotes low.

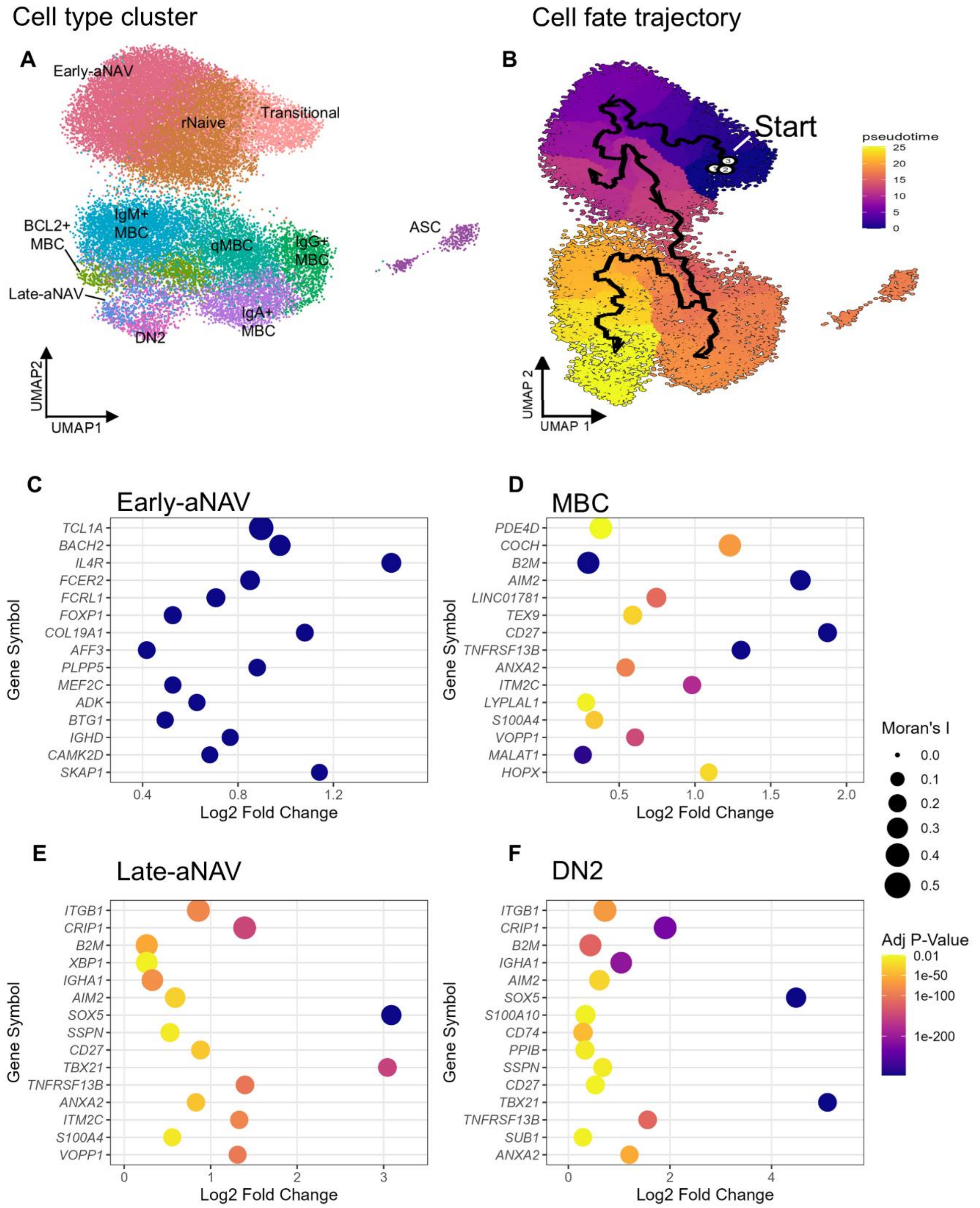


**Supplemental Figure 5. (A)** UMAP visualization of single-cell B cell populations (identical to Figure 3A), showing distinct clusters including transitional B cells, early-activated naïve (Early-aNAV) B cells, memory B cells (MBCs), late-activated naïve (Late-aNAV) B cells, and double-negative 2 (DN2) B cells. **(B)** Trajectory analysis depicting B cell differentiation dynamics using pseudotime ordering. The analysis was initiated with transitional B cells defined as the root/starting population (start node). Pseudotime represents an inferred temporal ordering of cells based on their transcriptional similarity, positioning cells along a continuous developmental trajectory independent of actual collection time. Cells are ordered from early (transitional) to more differentiated states, with branching paths illustrating divergent differentiation routes. The trajectory inference algorithm computes the most likely progression paths through transcriptional state space, with pseudotime values representing relative developmental progression. **(C-F)** Heatmaps showing the top 15 genes ranked by Moran's I spatial autocorrelation statistic for **(C)** Early-aNAV B cells, **(D)** Memory B cells (MBCs), **(E)** Late-aNAV B cells, and **(F)** DN2 B cells. Moran's I is a measure of spatial autocorrelation that identifies genes whose expression patterns exhibit non-random spatial organization along the pseudotime trajectory. Positive Moran's I values indicate that cells with similar expression levels for a given gene are located close together in pseudotime, suggesting coordinated regulation during differentiation. Genes with high Moran's I scores show smoothly varying expression patterns along the trajectory and represent key transcriptional signatures defining each B cell subset's position and transition within the developmental continuum.

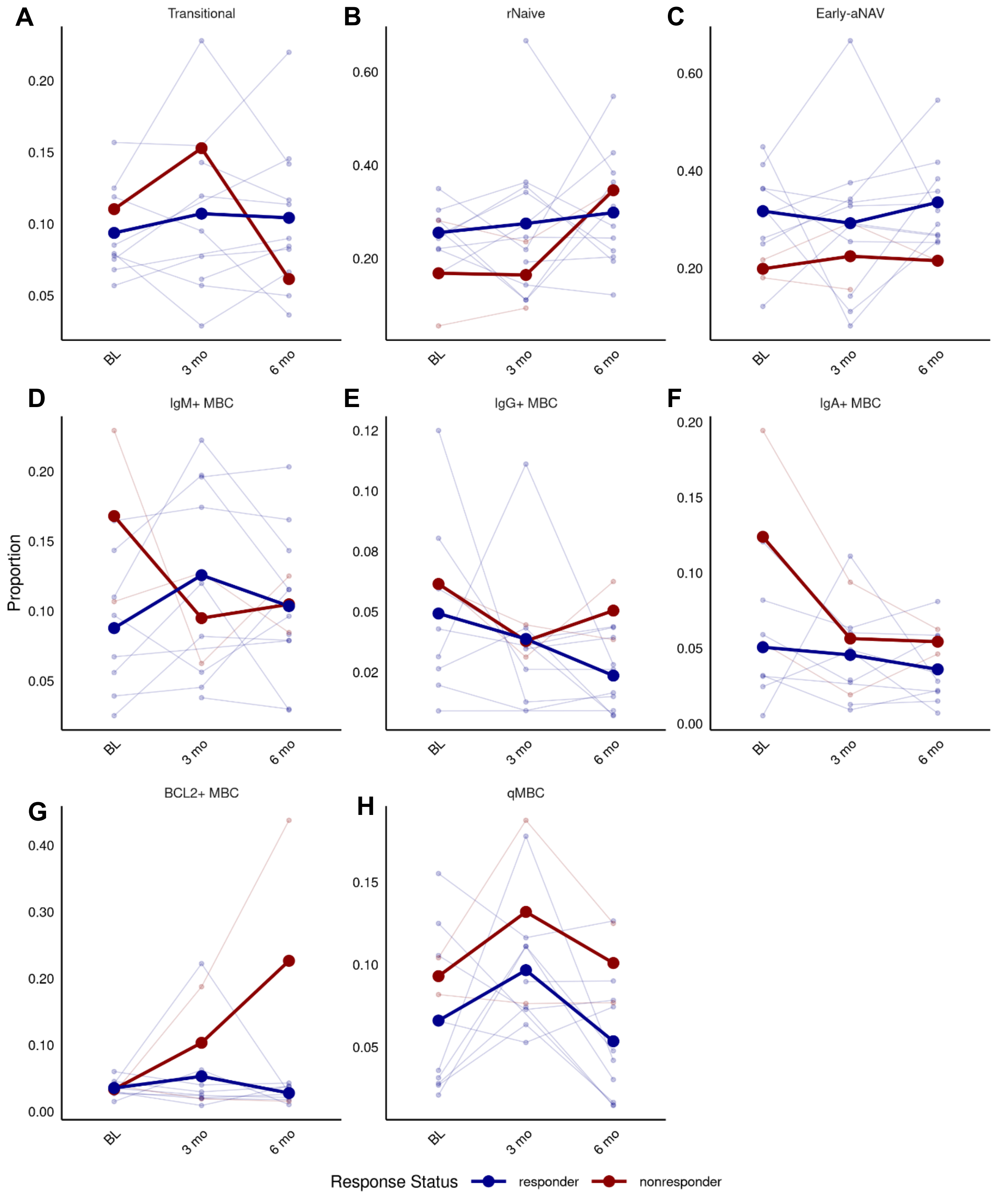


**Supplemental Figure 6.** Mean proportion of B cell subsets over time, related to Figure 4A-D. Proportion of **(A)** transitional, **(B)** rNaive, **(C)** early-aNAV, **(D)** IgM+ MBC, **(E)** IgG+ MBC, **(F)** IgA+ MBC, **(G)** BCL2+ MBC, and **(H)** qMBC at baseline (BL), 3 mos, and 6 mos. No significant differences were identified by chi-square comparing cell type proportions between responders and non-responders at 6 months.

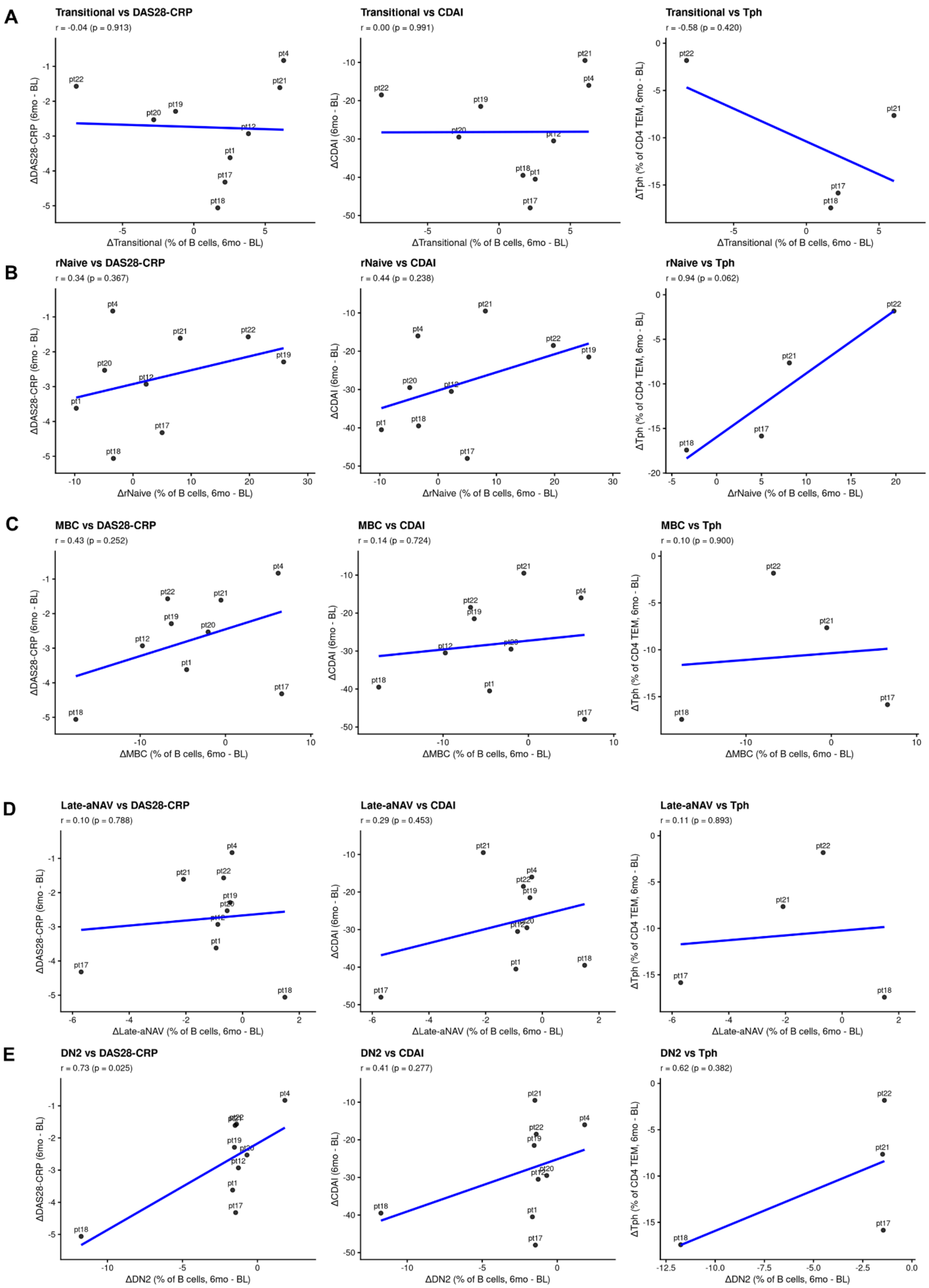

A
Transitional vs DAS28-CRP
r = -0.04 (p = 0.913)
ΔDAS28-CRP (6mo - BL)
ΔTransitional (% of B cells, 6mo - BL)
Transitional vs CDAI
r = 0.00 (p = 0.991)
ΔCDAI (6mo - BL)
ΔTransitional (% of B cells, 6mo - BL)
Transitional vs Tph
r = -0.58 (p = 0.420)
ΔTph (% of CD4 TEM, 6mo - BL)
ΔTransitional (% of B cells, 6mo - BL)
B
rNaive vs DAS28-CRP
r = 0.34 (p = 0.367)
ΔDAS28-CRP (6mo - BL)
ΔrNaive (% of B cells, 6mo - BL)
rNaive vs CDAI
r = 0.44 (p = 0.238)
ΔCDAI (6mo - BL)
ΔrNaive (% of B cells, 6mo - BL)
rNaive vs Tph
r = 0.94 (p = 0.062)
ΔTph (% of CD4 TEM, 6mo - BL)
ΔrNaive (% of B cells, 6mo - BL)
C
MBC vs DAS28-CRP
r = 0.43 (p = 0.252)
ΔDAS28-CRP (6mo - BL)
ΔMBC (% of B cells, 6mo - BL)
MBC vs CDAI
r = 0.14 (p = 0.724)
ΔCDAI (6mo - BL)
ΔMBC (% of B cells, 6mo - BL)
MBC vs Tph
r = 0.10 (p = 0.900)
ΔTph (% of CD4 TEM, 6mo - BL)
ΔMBC (% of B cells, 6mo - BL)
D
Late-aNAV vs DAS28-CRP
r = 0.10 (p = 0.788)
ΔDAS28-CRP (6mo - BL)
ΔLate-aNAV (% of B cells, 6mo - BL)
Late-aNAV vs CDAI
r = 0.29 (p = 0.453)
ΔCDAI (6mo - BL)
ΔLate-aNAV (% of B cells, 6mo - BL)
Late-aNAV vs Tph
r = 0.11 (p = 0.893)
ΔTph (% of CD4 TEM, 6mo - BL)
ΔLate-aNAV (% of B cells, 6mo - BL)
E
DN2 vs DAS28-CRP
r = 0.73 (p = 0.025)
ΔDAS28-CRP (6mo - BL)
ΔDN2 (% of B cells, 6mo - BL)
DN2 vs CDAI
r = 0.41 (p = 0.277)
ΔCDAI (6mo - BL)
ΔDN2 (% of B cells, 6mo - BL)
DN2 vs Tph
r = 0.62 (p = 0.382)
ΔTph (% of CD4 TEM, 6mo - BL)
ΔDN2 (% of B cells, 6mo - BL)

F

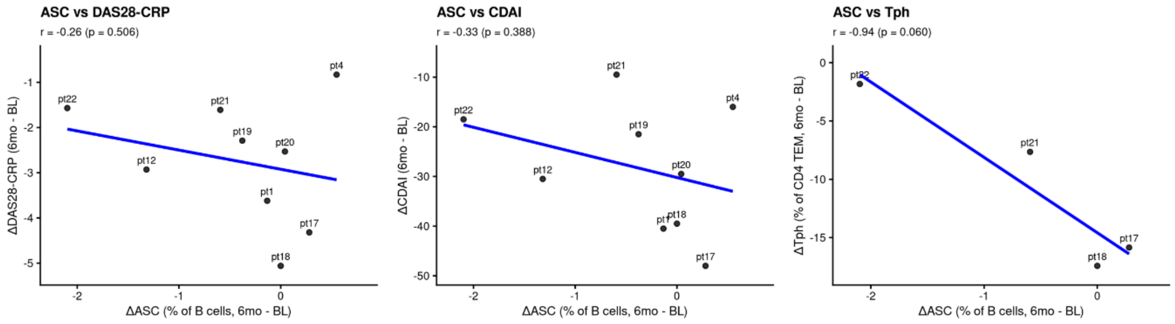


G

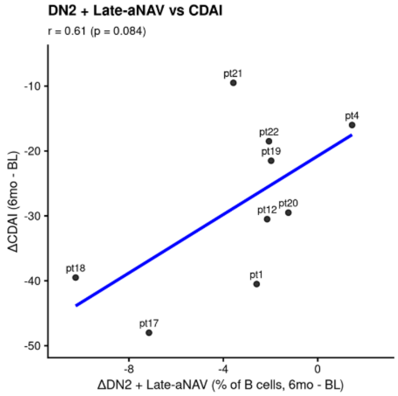


**Supplemental Figure 7.** Changes in Late-aNAV and DN2 B cell abundance correlate with clinical outcomes and peripheral helper T cell dynamics. Linear regression analyses examining the relationship between changes in B cell subset proportions and clinical/immunological parameters over the treatment period (baseline to 6 months). Each data point represents an individual patient, labeled with patient identifier. Change in **(A)** transitional B cells, **(B)** rNaive B cells, **(C)** Memory B cells, **(D)** late-aNAV, **(E)** DN2, and **(F)** ASCs compared to **(left)** Disease Activity Score 28 with C-reactive protein (ΔDAS28-CRP from baseline, 6mo - BL), **(middle)** Clinical Disease Activity Index (ΔCDAI, 6mo - BL), and **(right)** change in peripheral helper T cell (Tph) frequency (ΔTph % of CD4+ T cells, 6mo - BL). Tph cells provide help to B cells in peripheral tissues and contribute to autoantibody production in autoimmune disease. Blue lines represent linear regression fits. **(G)** Change in combination of DN2 + Late-aNAV cells vs. Clinical Disease Activity Index (ΔCDAI, 6mo - BL). Comparison of DN2 + Late-aNAV against DAS28-CRP and TPH are shown in Figure 4E and 4F. Correlation strength is assessed by Pearson correlation coefficient (r), with associated p-values indicating statistical significance. Negative changes in clinical scores (DAS28-CRP, CDAI) represent clinical improvement, while changes in cellular proportions can be positive or negative depending on expansion or contraction of each subset during treatment.

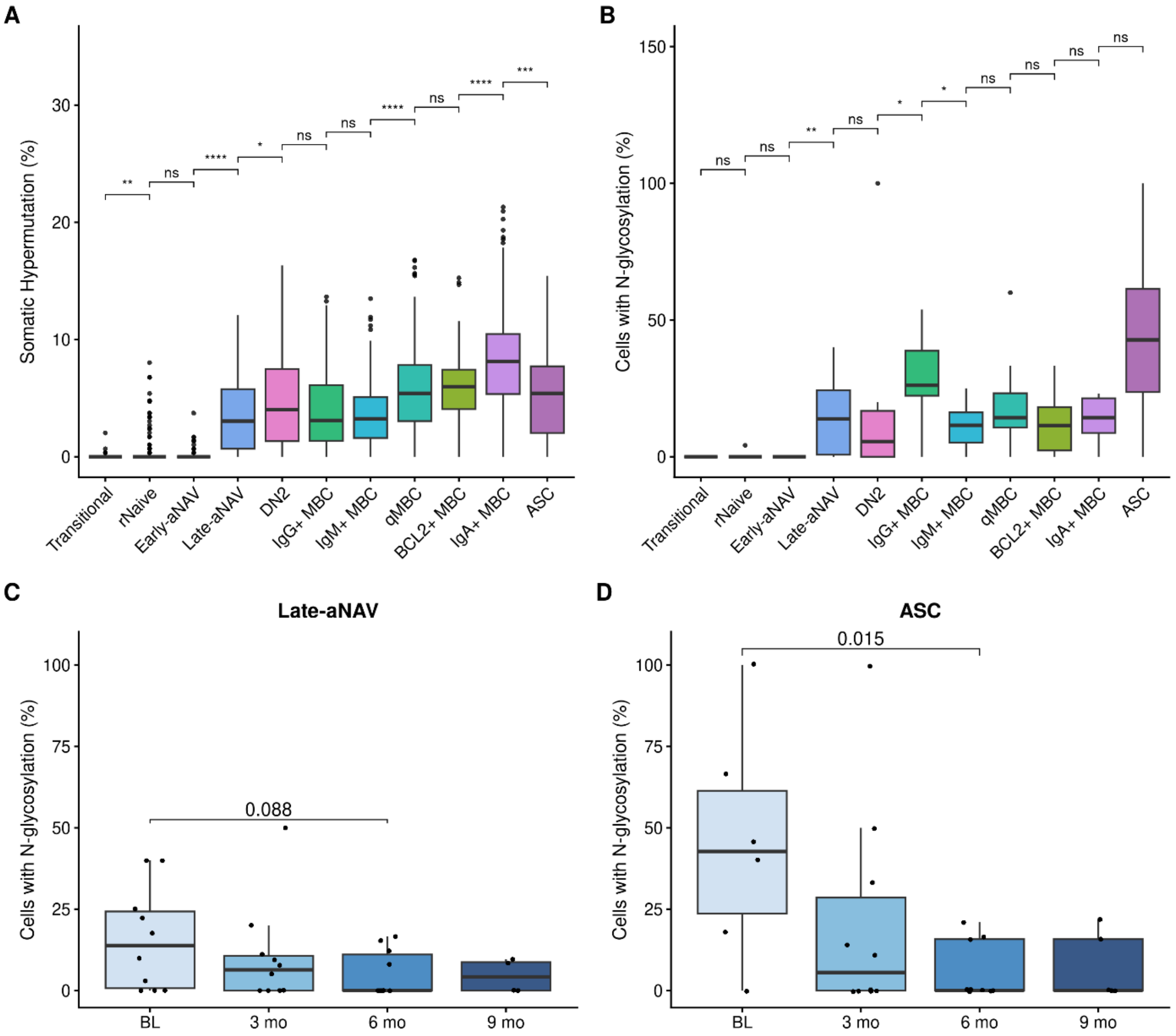


**Supplemental Figure 8. (A,B)** Somatic hypermutation (SHM) levels across B cell subsets at baseline. SHM is quantified as the percentage nucleotide sequence divergence of immunoglobulin heavy chain variable (IGHV) genes from their closest germline counterpart, as determined by IMGT alignment. Data encompass all B cells at the baseline timepoint, demonstrating the progressive accumulation of mutations from germline-proximal populations (Transitional and Naïve B cells) through intermediate states to highly mutated effector populations. The dashed line indicates germline sequence identity (0% SHM). **(A)** Displays SHM distribution across all major B cell subsets; **(B)** Presents the same data as percentage of sequences with N-glycosylation sites, highlighting functional consequences of accumulated mutations. **(C,D)** Prevalence of somatically-acquired N-linked glycosylation motifs in B cell receptors (BCRs). N-glycosylation motifs are defined by the canonical sequon: Asparagine-X-Serine/Threonine (N-X-S/T), where X represents any amino acid except Proline. To identify somatic variants, only sequences with evidence of SHM were analyzed (defined as <98% nucleotide identity to the closest germline IGHV gene by IMGT analysis), thereby excluding germline-encoded glycosylation sites. Data points represent the mean percentage of sequences per patient harboring non-germline N-glycosylation motifs. **(C)** Shows longitudinal dynamics in

Late-aNAV B cells across timepoints (baseline [BL], 3, 6, and 9 months post-treatment). **(D)** Displays N-glycosylation motif prevalence in antibody-secreting cells (ASCs) across the same timepoints. Box plots display the median (center line) and interquartile range (IQR; box boundaries); whiskers extend to 1.5× IQR, with outliers shown as individual open circles. Statistical comparisons between groups were performed using the non-parametric Wilcoxon rank-sum test. Significance levels: $^*p < 0.05$, $^{**}p < 0.01$, $^{***}p < 0.001$.

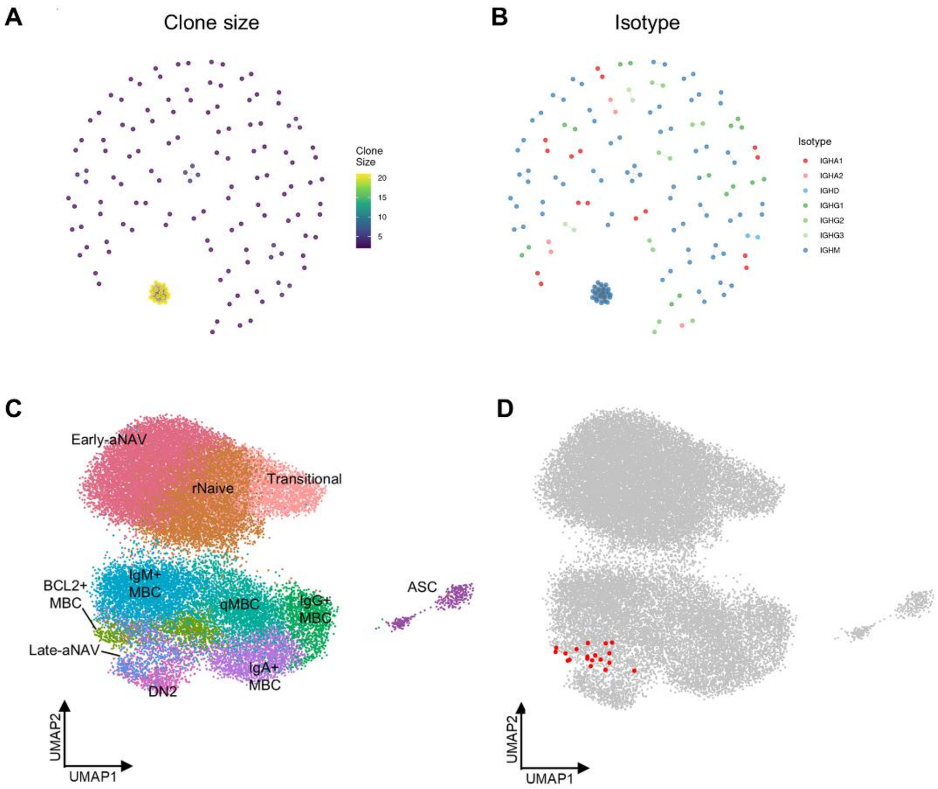


E

| celltype | Count | Percentage |
|---|---|---|
| BCL2+ MBC | 13 | 68.42 |
| DN2 | 4 | 21.05 |
| aNAV-Late | 2 | 10.53 |

**Supplemental Figure 9**. **(A,B)** Clone size and isotype distribution of B cell clones. The largest clone was defined as Clone 1 **(C)** UMAP visualization of B cell subsets (identical to Figure 3A), showing the transcriptional landscape of distinct B cell populations including Early-aNAV (early-activated naïve), Naïve, Transitional, BCL2+ memory B cells (MBCs), Late-aNAV (late-activated naïve), DN2 (double-negative 2), MBC (memory B cells), and antibody-secreting cells (ASCs). Each point represents an individual cell, colored by cluster identity based on gene expression profiles. **(D)** Spatial distribution of Clone 1 cells overlaid on the UMAP projection. Clone 1 represents a clonally expanded B cell lineage sharing identical immunoglobulin heavy and light chain V(D)J rearrangements (n=19 cells total). Cells belonging to Clone 1 are highlighted in red, while all other cells are shown in gray. **(E)** This clonotype exhibits multi-subset distribution across the B cell differentiation spectrum. The presence of a single clonotype across multiple transcriptionally distinct subsets indicates clonal persistence and differentiation of a common B cell ancestor through different functional states.

A

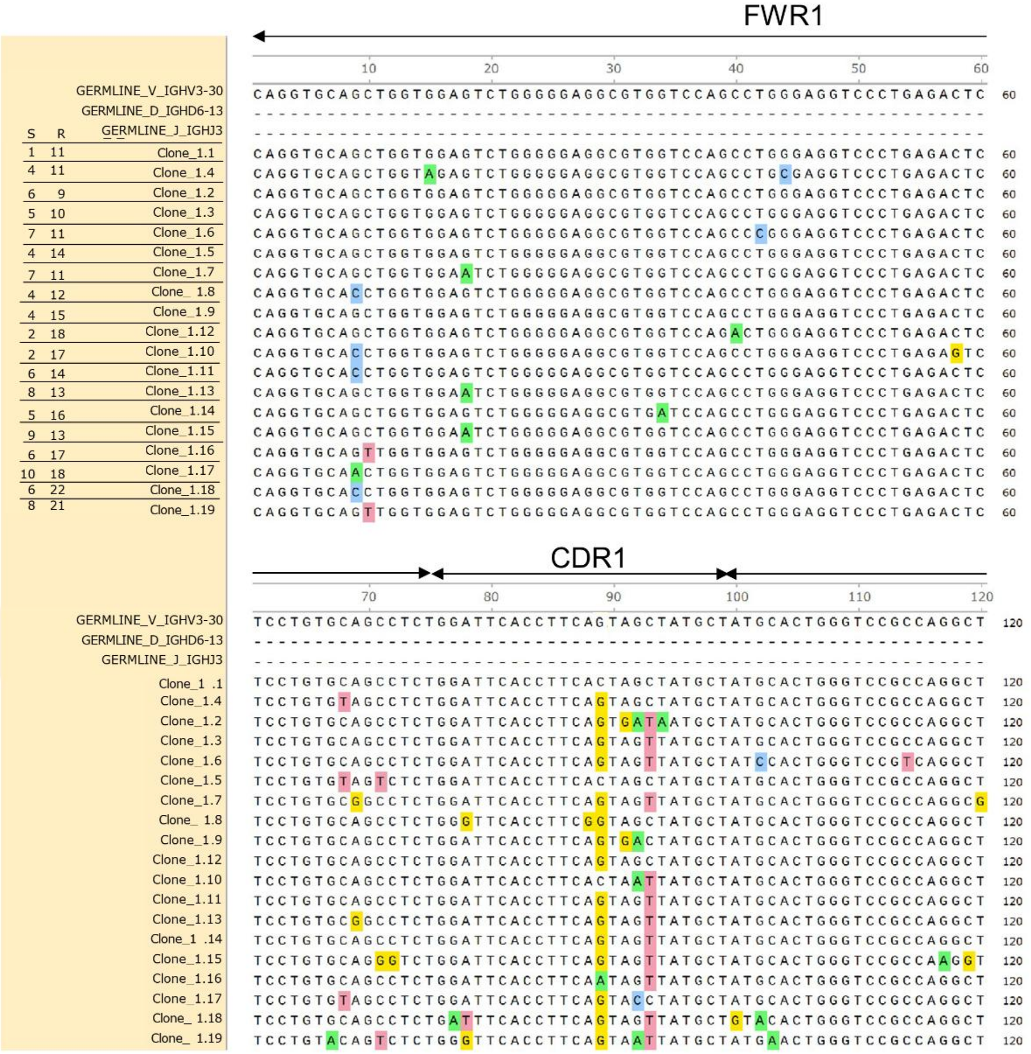
FWR1
CDR1
GERMLINE_V_IGHV3-30
GERMLINE_D_IGHD6-13
GERMLINE_J_IGHJ3
S R
1 11 Clone_1.1
4 11 Clone_1.4
6 9 Clone_1.2
5 10 Clone_1.3
7 11 Clone_1.6
4 14 Clone_1.5
7 11 Clone_1.7
4 12 Clone_ 1.8
4 15 Clone_1.9
2 18 Clone_1.12
2 17 Clone_1.10
6 14 Clone_1.11
8 13 Clone_1.13
5 16 Clone_1.14
9 13 Clone_1.15
6 17 Clone_1.16
10 18 Clone_1.17
6 22 Clone_1.18
8 21 Clone_1.19

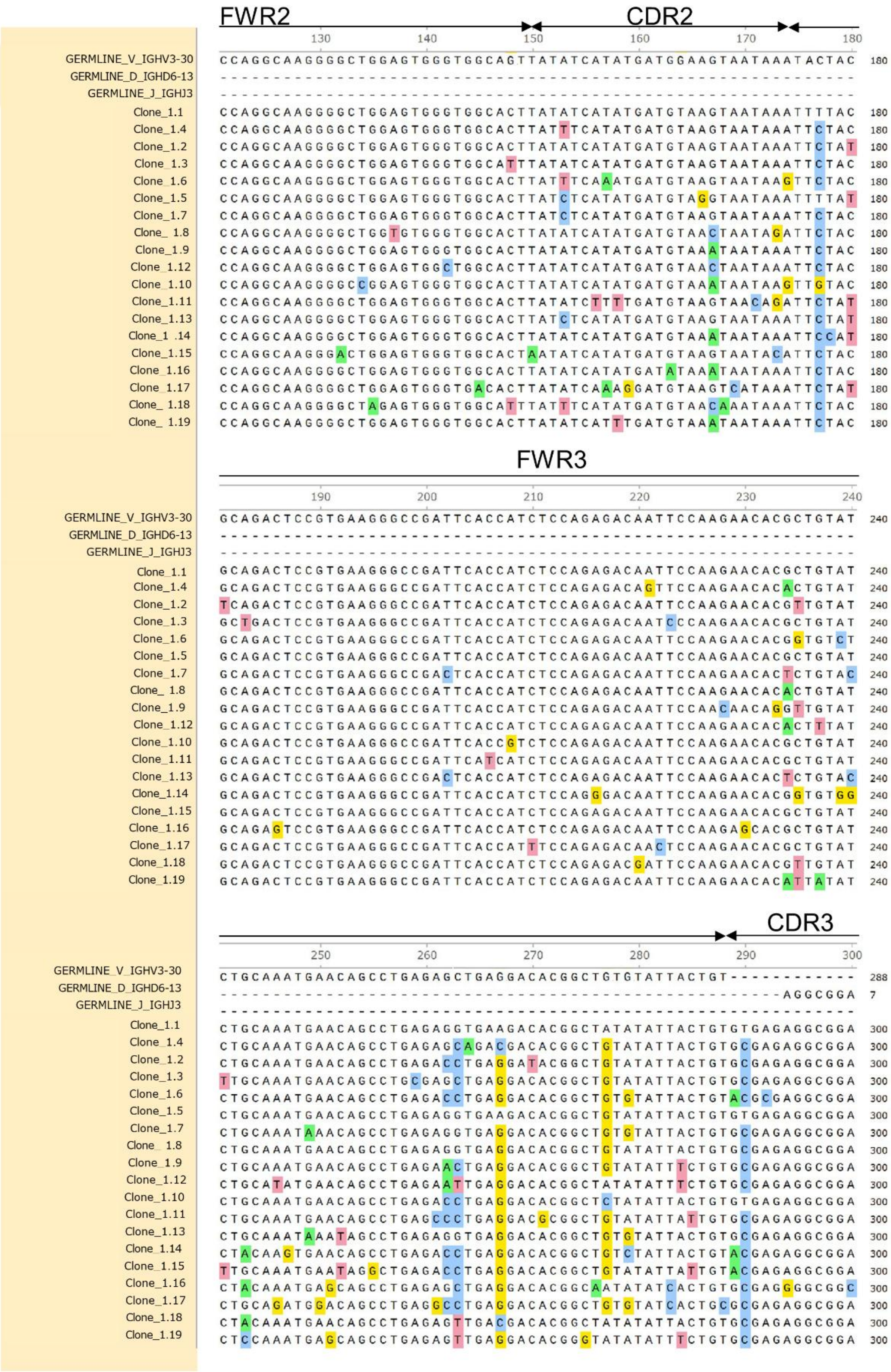
FWR2
CDR2
FWR3
CDR3
GERMLINE_V_IGHV3-30
GERMLINE_D_IGHD6-13
GERMLINE_J_IGHJ3
Clone_1.1
Clone_1.4
Clone_1.2
Clone_1.3
Clone_1.6
Clone_1.5
Clone_1.7
Clone_ 1.8
Clone_1.9
Clone_1.12
Clone_1.10
Clone_1.11
Clone_1.13
Clone_1 .14
Clone_1.15
Clone_1.16
Clone_1.17
Clone_ 1.18
Clone_ 1.19

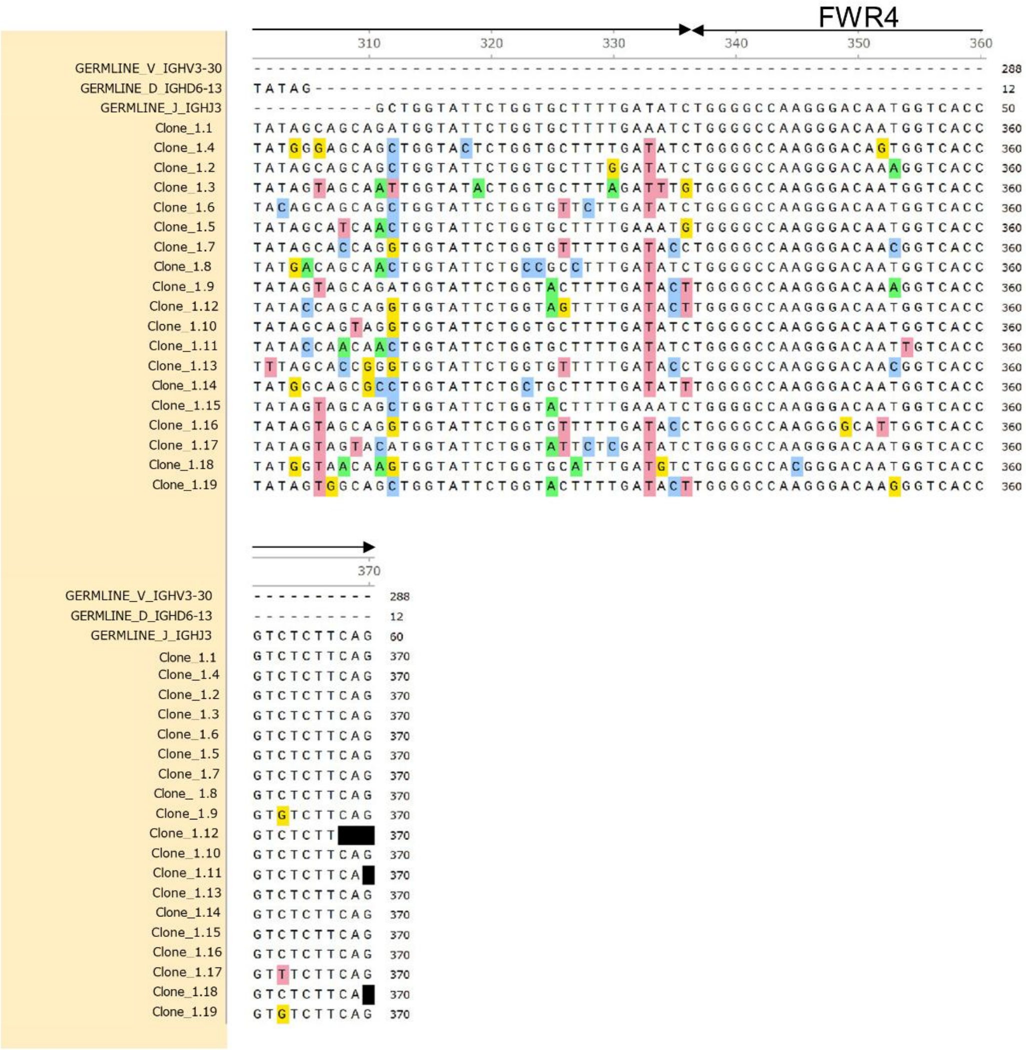
FWR4
310
320
330
340
350
360
370
GERMLINE_V_IGHV3-30
GERMLINE_D_IGHD6-13
GERMLINE_J_IGHJ3
Clone_1.1
Clone_1.4
Clone_1.2
Clone_1.3
Clone_1.6
Clone_1.5
Clone_1.7
Clone_1.8
Clone_1.9
Clone_1.12
Clone_1.10
Clone_1.11
Clone_1.13
Clone_1.14
Clone_1.15
Clone_1.16
Clone_1.17
Clone_1.18
Clone_1.19

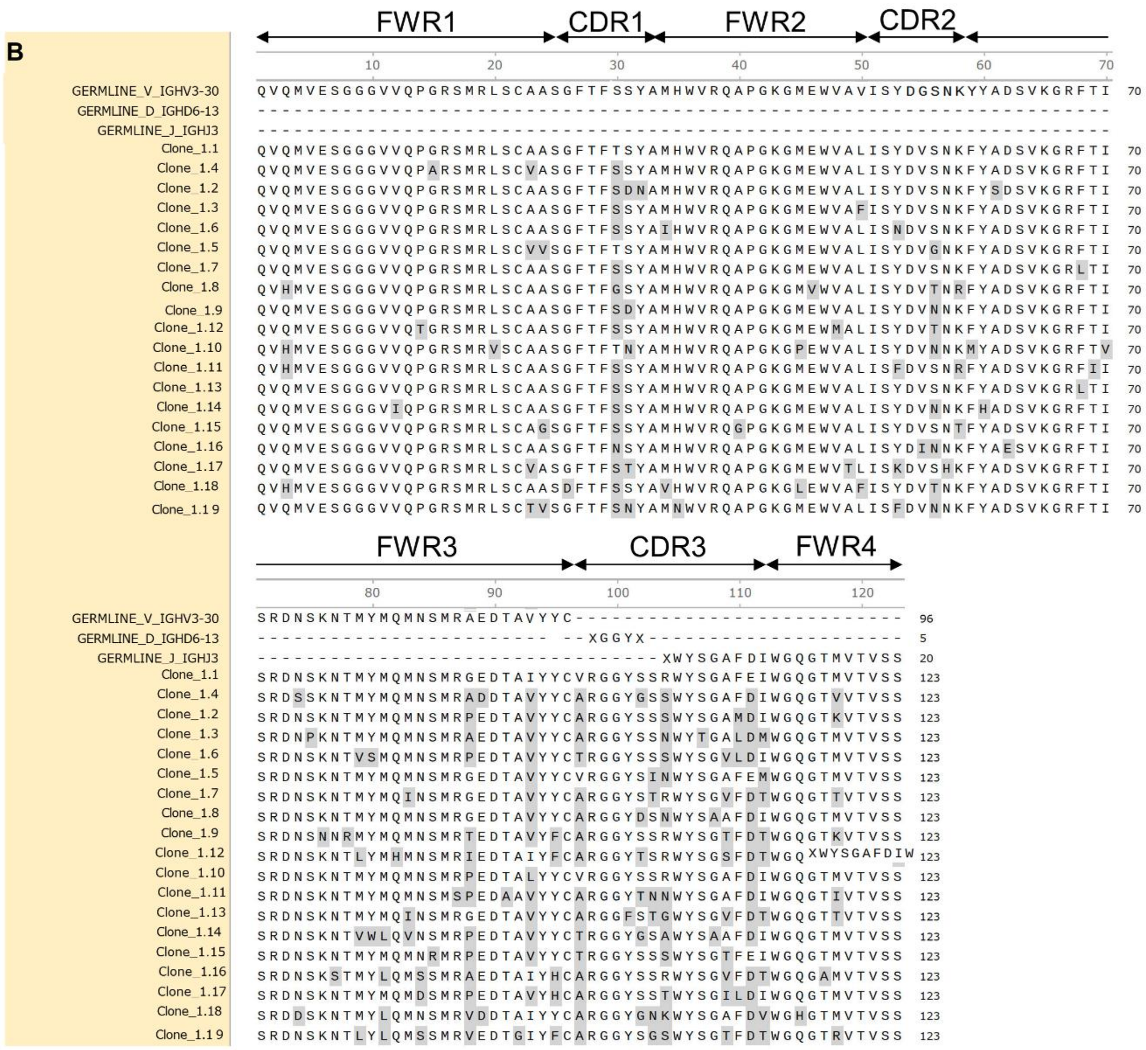

**Supplemental Figure 10.** Nucleotide and amino acid sequence alignment of Clone 1 B cell receptor (BCR) heavy chain sequences. **(A)** Nucleotide sequence alignment of 19 clonally related BCR heavy chain sequences (Clone 1.1-1.19) aligned against the predicted germline V (IGHV3-30), D (IGHD6-13), and J (IGHJ3) gene segments. Silent (S) and replacement (R) mutations compared to germline are noted. **(B)** Corresponding amino acid sequence alignment of the same 19 clonal members. Germline V, D, and J gene segments are displayed at the top of each alignment as reference sequences. Immunoglobulin region boundaries — Framework regions (FWR1–FWR4) and Complementarity-Determining Regions (CDR1–CDR3) — are annotated according to the IMGT numbering system (Lefranc et al., 2003), where FWR3 terminates at the conserved Cysteine (C104, encoded by TGT) and CDR3 begins immediately thereafter, encompassing N-nucleotide additions, the D-segment, and extending through to the conserved Tryptophan (W118, encoded by TGG) at the 5' end of the J-segment, after which FWR4 begins. Clone 1.1 serves as the reference sequence for variant highlighting; nucleotide bases (A) or amino acid residues (B) that differ from Clone 1.1 are marked with four-color highlighting, where each color corresponds to a specific base or residue identity. Identical positions are unmarked. Clonal daughter sequences (Clone 1.1–1.19) are ordered based on physicochemical properties and mutual sequence similarity. Germline gene assignments and

CDR/FWR annotations were determined using IgBLAST against the IMGT reference directory. All alignments were generated and visualized using SnapGene. The somatic hypermutation pattern across clonal daughters reflects intraclonal diversification consistent with ongoing affinity maturation within the germinal center.

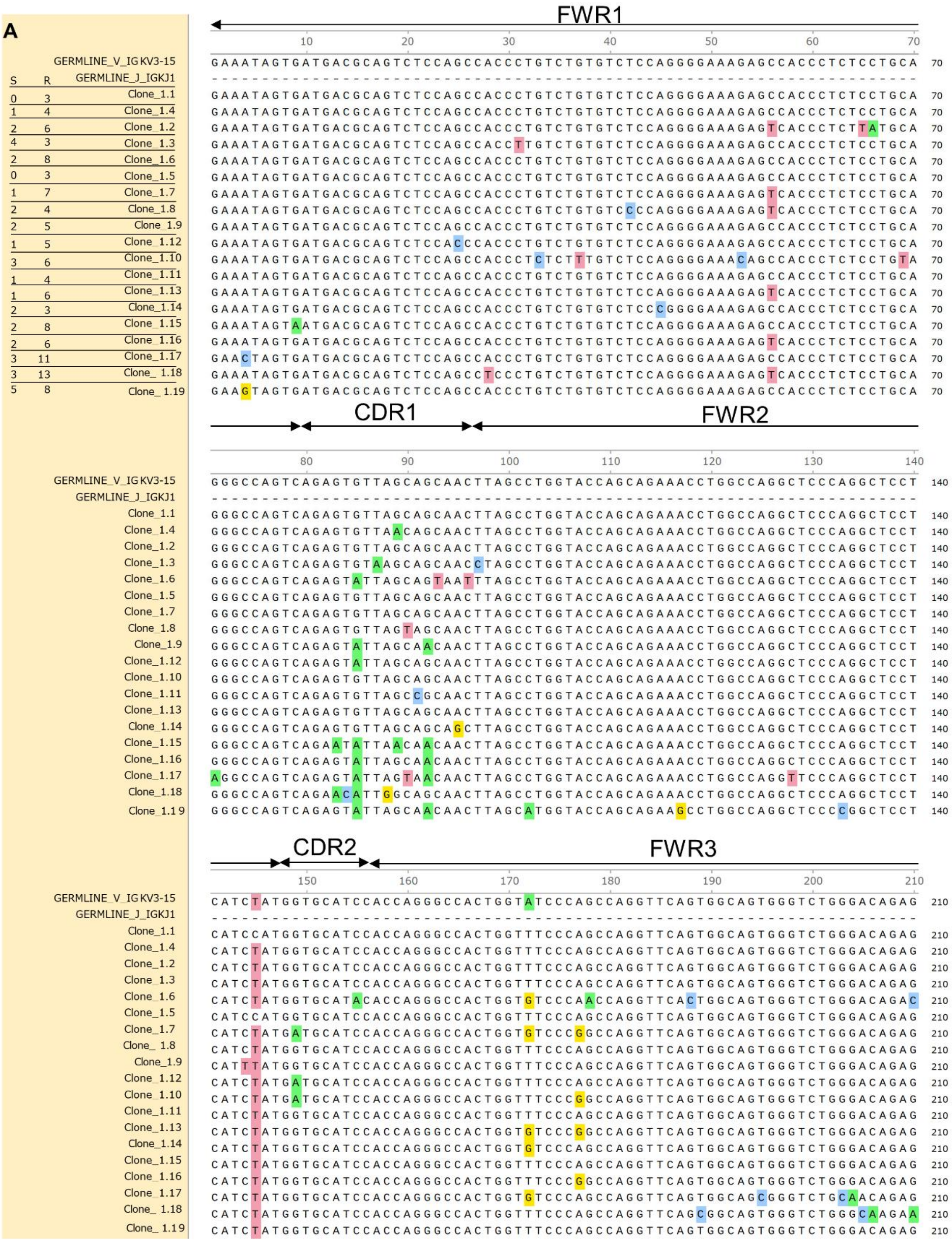
A
FWR1
CDR1
FWR2
CDR2
FWR3
GERMLINE_V_IGKV3-15
GERMLINE_J_IGKJ1
S R
0 3 Clone_1.1
1 4 Clone_1.4
2 6 Clone_1.2
4 3 Clone_1.3
2 8 Clone_1.6
0 3 Clone_1.5
1 7 Clone_1.7
2 4 Clone_1.8
2 5 Clone_1.9
1 5 Clone_1.12
3 6 Clone_1.10
1 4 Clone_1.11
1 6 Clone_1.13
2 3 Clone_1.14
2 8 Clone_1.15
2 6 Clone_1.16
3 11 Clone_1.17
3 13 Clone_1.18
5 8 Clone_1.19

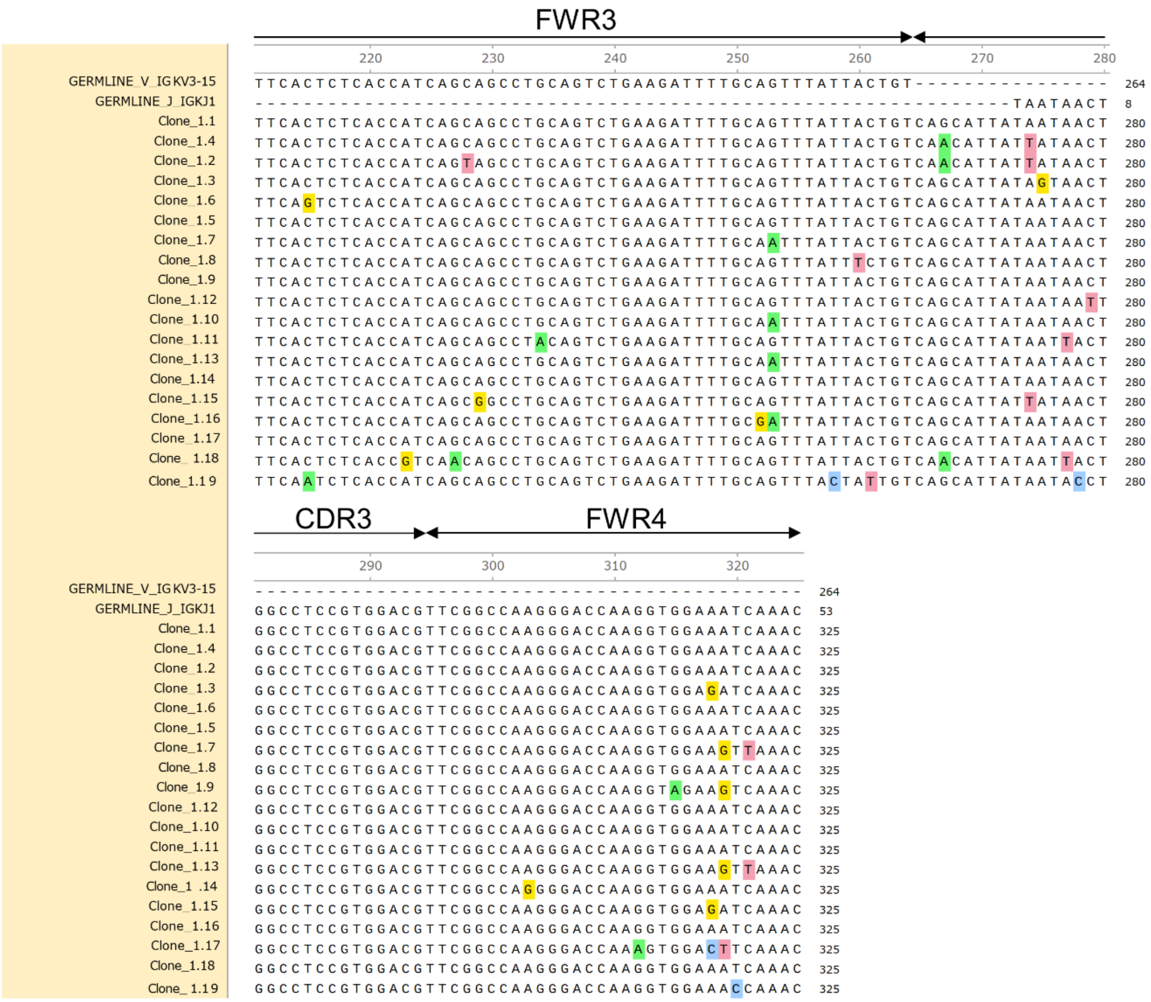
FWR3
220
230
240
250
260
270
280
GERMLINE_V_IGKV3-15
GERMLINE_J_IGKJ1
Clone_1.1
Clone_1.4
Clone_1.2
Clone_1.3
Clone_1.6
Clone_1.5
Clone_1.7
Clone_1.8
Clone_1.9
Clone_1.12
Clone_1.10
Clone_1.11
Clone_1.13
Clone_1.14
Clone_1.15
Clone_1.16
Clone_1.17
Clone_1.18
Clone_1.19
CDR3
FWR4
290
300
310
320

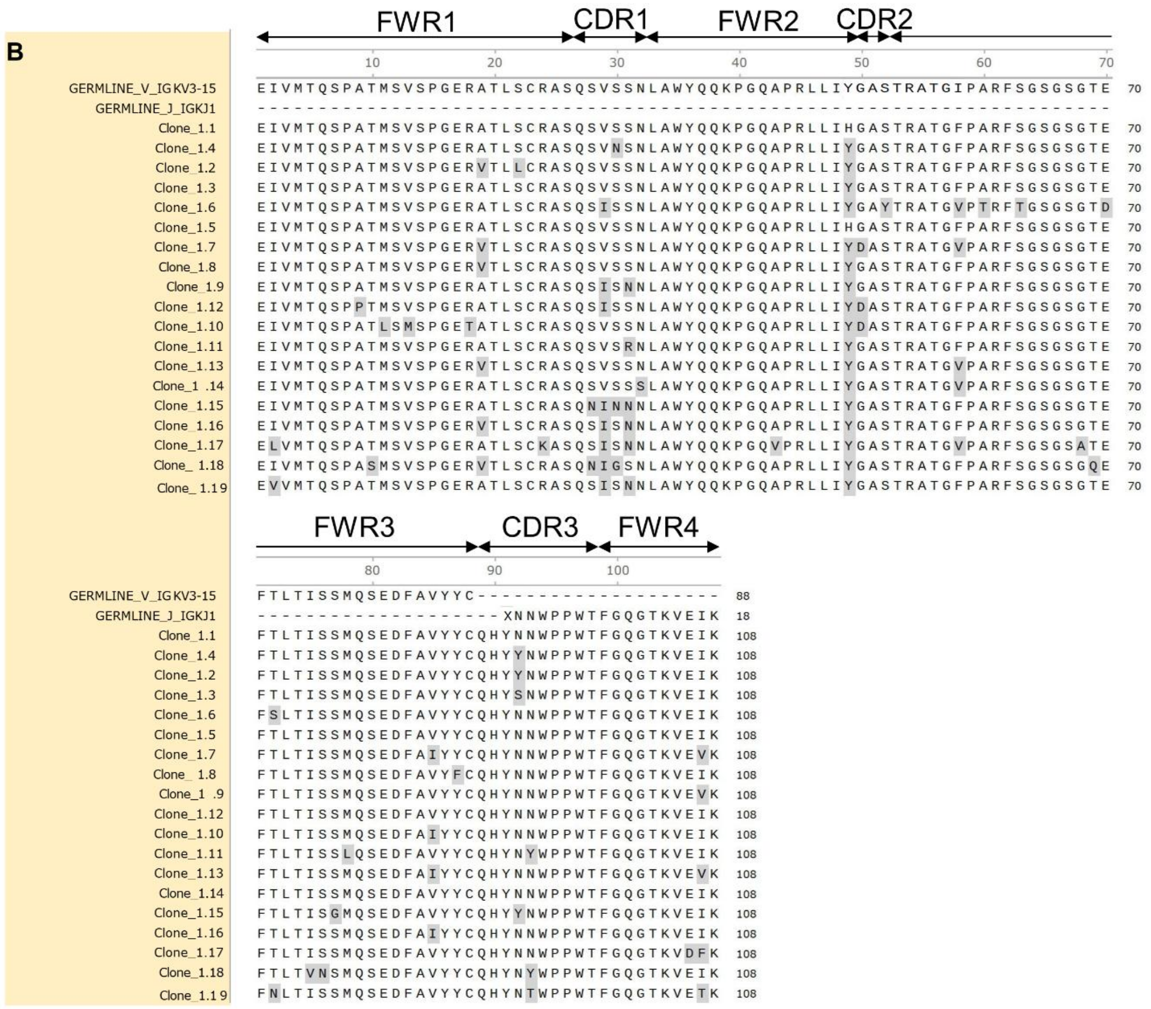

**Supplemental Figure 11.** Nucleotide and amino acid sequence alignment of Clone 1 B cell receptor (BCR) light chain (Igκ) sequences. **(A)** Nucleotide sequence alignment of 19 clonally related BCR immunoglobulin kappa (Igκ) light chain sequences (Clone 1.1–1.19) aligned against the predicted germline V (IGKV3-15) and J (IGKJ1) gene segments. Silent (S) and replacement (R) mutations compared to germline are noted. **(B)** Corresponding amino acid sequence alignment of the same 19 clonal members. Germline V and J gene segments are displayed at the top of each alignment as reference sequences. Note that light chain immunoglobulin genes do not utilize a Diversity (D) gene segment, and therefore the VJ junction is direct, with any non-templated nucleotides representing N-additions introduced during V(D)J recombination. Immunoglobulin region boundaries — Framework regions (FWR1–FWR4) and Complementarity-Determining Regions (CDR1–CDR3) — are annotated according to the IMGT numbering system (Lefranc et al., 2003), where FWR3 terminates at the conserved Cysteine (C104) and CDR3 begins immediately thereafter, extending through to the conserved Phenylalanine (F118) at the 5' end of the J-segment, after which FWR4 begins. Clone 1.1 serves as the reference sequence for variant highlighting; nucleotide bases (A) or amino acid residues (B) that differ from Clone 1.1 are marked with four-color highlighting, where each color corresponds to a specific base or residue identity. Identical positions are unmarked. Clonal daughter sequences (Clone 1.1–1.19) are ordered based on physicochemical properties and mutual sequence similarity. Germline gene assignments and CDR/FWR annotations were determined using IgBLAST against the IMGT reference directory. All alignments were generated and visualized using SnapGene. The somatic hypermutation pattern across clonal daughters reflects intraclonal diversification of both heavy and light chains, consistent with ongoing affinity maturation within the germinal center.

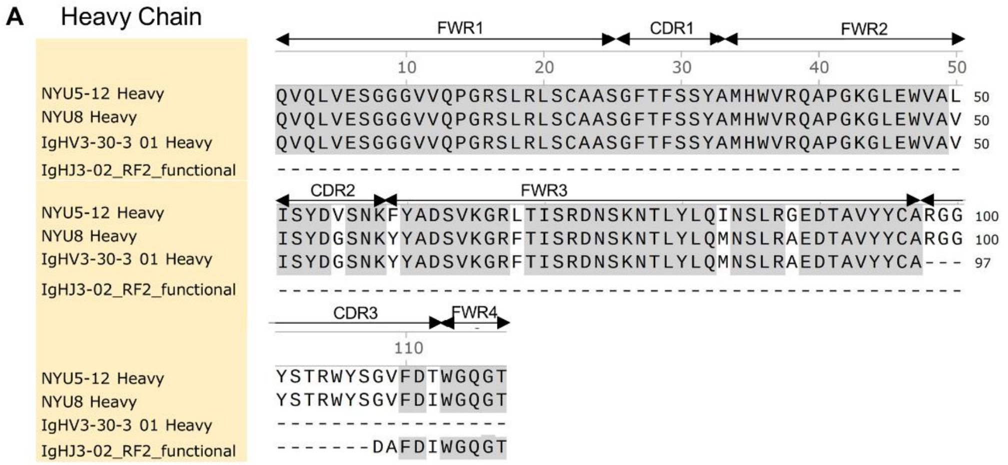


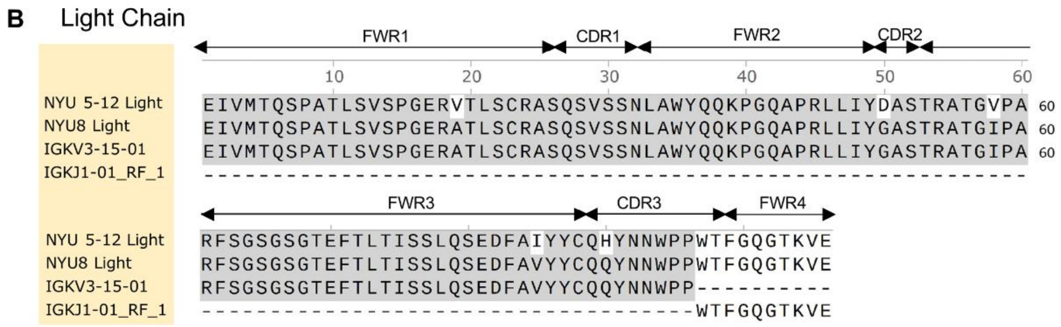


**Supplemental Figure 12**. Sequence of **(A)** heavy chain and **(B)** light chain recombinant antibody produced (NYU5-12; Clonotype 1 from Figure 3), and germline reverted antibody (NYU8) produced, aligned to closest germline sequence. RF represents reading frame used for alignment. Aligned using Clustal Omega.

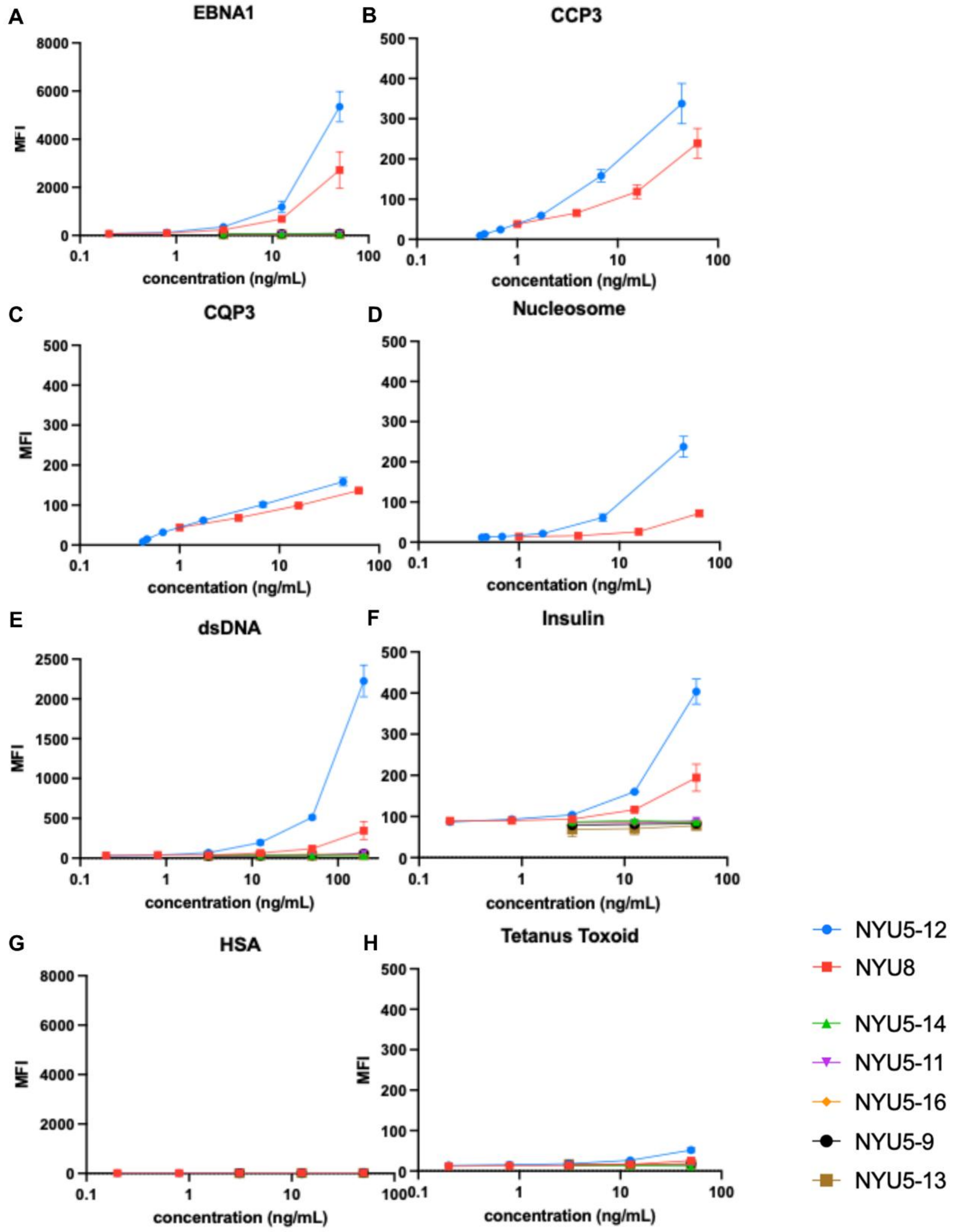


**Supplemental Figure 13.** Antigen binding specificity of recombinant monoclonal antibody NYU5-12 (Clone 1) and germline-reverted antibody NYU8. Binding reactivity of recombinant monoclonal antibodies derived from a patient with rheumatoid arthritis (RA) was assessed by multiplex bead-based immunoassay (MagPix/Luminex). Results are reported as Net Median Fluorescence Intensity (Net MFI). NYU5-12 is the somatically hypermutated monoclonal antibody produced from the dominant expanded clonotype (Clone 1). NYU8 is the corresponding germline-reverted antibody, in which somatic hypermutations in both the heavy chain (IGHV3-30) and light chain (IGKV3-15) were reverted to their predicted germline sequences to assess the contribution of affinity maturation to antigen specificity. NYU5-12 and NYU 8 binding to **(A)** EBNA1, **(B)** CCP-3, **(C)** CQP3 (the corresponding native arginine-containing control peptide to CCP3), **(D)** Nucleosome, **(E)** dsDNA, **(F)** Insulin, **(G)** human serum albumin, and **(H)** Tetanus Toxoid. Additional recombinant monoclonal antibodies derived from the same RA patient (NYU 5-14, 5-11, 5-16, 5-9, and 5-13 are unrelated and did not show binding to any antigens tested) did not demonstrate any specific reactivity.
